# Comparative analysis of White and African American groups reveals unique lipid and inflammatory features of diabetes

**DOI:** 10.1101/2024.11.13.24317202

**Authors:** Gabriela Pacheco Sanchez, Miranda Lopez, Leandro M. Velez, Ian Tamburini, Naveena Ujagar, Julio Ayala, Gabriela De Robles, Hannah Choi, John Arriola, Rubina Kapadia, Alan B. Zonderman, Michele K. Evans, Cholsoon Jang, Marcus M. Seldin, Dequina A. Nicholas

**Author notes:** Corresponding author: Dequina Nicholas. These authors contributed equally (Gabriela Pacheco Sanchez, Miranda Lopez).

## Abstract

**Importance:** African Americans have a higher prevalence of Type 2 Diabetes (T2D) compared to White groups. T2D is a health disparity clinically characterized by dysregulation of lipids and chronic inflammation. However, how the relationships among biological and sociological predictors of T2D drive this disparity remains to be addressed.

**Objective:** To determine characteristic plasma lipids and systemic inflammatory biomarkers contributing to diabetes presentation between White and African American groups.

**Design:** We performed a cross-sectional retrospective cohort study using pre-existing demographic and clinical data from two diverse studies: Healthy Aging in Neighborhoods of Diversity across the Life Span (HANDLS) and AllofUs. From HANDLS (N=40), we used information from wave 1 (2004). From AllofUs (N=17,339), we used data from the Registered Tier Dataset v7, available in the AllofUs researcher workbench.

**Setting:** HANDLS is a population-based cohort study involving 3720 participants in the Baltimore area supported by the Intramural Research Program of the National Institute on Aging. HANDLS is a longitudinal study designed to understand the sources of persistent health disparities in overall longevity and chronic disease in White and African American individuals. The AllofUs study is an NIH funded multicenter study consisting of patient-level data from 331,382 individuals from 35 hospitals in the United States aimed at sampling one million or more people living in the United States to provide a collection of broadly accessible data.

**Participants:** The HANDLS subcohort participants (N=40) were divided into four groups equally distributed by race, sex, and diabetes status. Groups were also matched by age, body mass index, and poverty status. The analysis pipeline consisted of evaluating the significance of the variables race and disease status using the 2-way ANOVA test and post-ANOVA comparisons using Fisher LSD test, reporting unadjusted p-values. Additionally, unsupervised (PCA) and supervised (OPLS-DA) clustering analysis was performed to determine putative biological drivers of variability and main immunological and metabolic features characterizing diabetes in White and African American groups from HANDLS. Major clinical findings were validated in a large cohort of White and African American groups with T2D in the AllofUS research study (N=17,339). AllofUs groups were of similar range in age and BMI as HANDLS. Furthermore, a linear regression model was built adjusting for age and BMI to determine differences in clinical findings between White and African American groups with T2D.

**Main Outcomes and Measures:** Primary outcomes using a HANDLS subcohort (N=40) were clinical parameters related to diabetes, plasma lipids determined by lipidomics and measured by mass spectrometry, and cytokine profiling using a customized panel of 52 cytokines and growth factors measured by Luminex. Outcomes evaluated in the AllofUs study (N=17,339) were clinical: cholesterol to HDL ratio, triglycerides, fasting glucose, insulin, and hemoglobin A1C.

**Results:** In the HANDLS subcohort, White individuals with diabetes had elevated cholesterol to HDL ratio (mean difference -1.869, *p*=0.0053), high-sensitivity C-reactive protein (mean difference -9.135, *p*=0.0040), and clusters of systemic triglycerides measured by lipidomics, compared to White individuals without diabetes. These clinical markers of dyslipidemia (cholesterol to HDL ratio and triglycerides) and inflammation (hs-CRP) were not significantly elevated in diabetes in African Americans from the HANDLS subcohort. These results persisted even when controlling for statin use. Diabetes in White individuals in the HANDLS cohort was characterized by a marked elevation in plasma lipids, while an inflammatory status characterized by Th17-cytokines was predominant in the African American group from the HANDLS subcohort. We validated the key findings of elevated triglycerides and cholesterol to HDL ratio in White individuals with T2D in a sample (N=17,339) of the AllofUs study.

**Conclusions and Relevance:** Our results show that diabetes can manifest with healthy lipid profiles, particularly in these cohorts of African Americans. This study suggests that Th17-inflammation associated with diabetes is characteristic of African Americans, while a more classic inflammation is distinctive of White individuals from HANDLS cohort. Further, clinical markers of dyslipidemia seem to characterize diabetes presentation only in White groups, and not in African Americans.

## INTRODUCTION

Diabetes affects a staggering 38.4 million individuals in the United States which is an estimated 11.6% of the entire population (Centers for Disease Control and Prevention (CDC), 2024). Diabetes is an impactful health disparity given its disproportionate incidence in some populations from different ethnic and racial backgrounds (Rodríguez & Campbell, 2017). Approximately 15 million diagnosed and undiagnosed people with diabetes belong to minority health populations, including non-Hispanic African American, non-Hispanic Asian, and Hispanic groups. People from these populations bear significant proportions of the burden and complications of diabetes within the US (Centers for Disease Control and Prevention (CDC), 2024; Haw et al., 2021).

Insulin resistance and clinical measurements of glucose management like HbA1C, blood insulin, and blood glucose are preferably used in the study of diabetes (Christensen et al., 2010; Khalili et al., 2023; Khan et al., 2018). However, presentation of diabetes is also characterized by dysregulation of lipid metabolism (dyslipidemia) and chronic inflammation (Calle & Fernandez, 2012; Galicia-Garcia et al., 2020; Mooradian, 2009). Efforts to study diabetes-related metabolic phenotypes in diverse populations have provided insights into the characterization of lipid profiles and inflammatory markers in populations with diabetes. For example, Yang and collaborators (Yang et al., 2023) found that Chinese patients with prediabetes and diabetes had an increased abundance of plasma ceramides including, Cer(d18:1/22:0), Cer(d18:1/23:0), and Cer(d18:1/24:0) compared to healthy Chinese control patients, revealing a novel relationship between diabetes and plasma lipid profiles previously unappreciated. Regarding the relationship of inflammation to diabetes, reports have historically found an increase in systemic markers such as C-reactive protein (CRP), and the pro-inflammatory cytokines interleukin 6 (IL-6) and tumor necrosis factor alpha (TNF-α) as features of diabetes (Popko et al., 2010; Stanimirovic et al., 2022; Swaroop et al., 2012). More recent literature in type 2 diabetes (T2D) research has discovered an elevation of Th17 cytokines (IL-17A, IL-17E, IL-17F, IL-21, and IL-22) in people with T2D. Particularly, IL-17A has been reported to be elevated in patients with T2D compared to patients without T2D (Ip et al., 2015, 2016; Nicholas et al., 2019). Notably, an interesting study demonstrated the potential association between Th17 inflammatory profiles and compromised lipid metabolism in the context of fatty acid uptake in T2D (Nicholas et al., 2019). However, these studies were not powered to assess the relationship between lipids and Th17 inflammation by race and ethnicity. Studies in the field of cardiometabolic disease provide evidence to support that disparities persist regarding clinical features of diabetes in diverse populations. For example, disparate levels of lipids (cholesterol, high-density lipoprotein (HDL), low-density lipoprotein (LDL), and triglycerides) have been reported in African Americans compared to White individuals in the context of cardiometabolic disease (Koutroumpakis et al., 2016; Riccardi et al., 2020).

Given the known health disparities in diabetes, our goal was to determine the relationship between lipid profiles and inflammatory markers in diabetes within White and African American cohorts. We hypothesized that the relationship of lipids to inflammation in the presentation of diabetes varies across diverse populations. To this end, we specifically compared clinical measures of dyslipidemia and inflammation and unbiased biochemical measures of plasma lipids and cytokines between White and African American individuals using multivariate analyses.

## RESEARCH DESIGN AND METHODS

### HANDLS Population

Study participants were sampled from the Healthy Aging in Neighborhoods of Diversity across the Life Span (HANDLS) (Evans et al., 2010), a population-based cohort study involving 3720 participants supported by the Intramural Research Program of the National Institute on Aging.

<u>HANDLS</u> is a longitudinal study designed to understand the sources of persistent health disparities in overall longevity and chronic disease in White and African American individuals. For this study, participants were recruited from 13 contiguous U.S. Census tracts in the Baltimore area and were 30 to 64 years of age at the time of enrollment. A subsample of 40 participants was randomly selected and divided into 4 matched comparison groups based on race and disease status (No diagnosis of diabetes=NoDx; diagnosis of diabetes=Dx). These groups are White individuals without diabetes (NoDx-White), White people with diabetes (Dx-White), African Americans without diabetes (NoDx-AA), and African Americans with diabetes (Dx-AA). Based on the absence of insulin use, 80% of the cohort with diabetes had confirmed T2D. To maximize the potential to identify different biological contributors to health disparities, the comparison groups were equally distributed by race, diabetes status, and sex, with each group being matched by age, body mass index (BMI), and poverty status (**Table 1**). Importantly, all statistical comparisons in this study were performed between matched groups, based on diabetes status and race. As such, we evaluated comparisons between NoDx-White vs Dx-White, NoDx-AA vs Dx-AA, NoDx-White vs NoDx-AA, and Dx-White vs Dx-AA.

**Table 1.** HANDLS subcohort demographics. **Table legend:** A subcohort of 40 individuals from the HANDLS study were divided into 4 comparison groups based on disease status and race: White without diabetes (NoDx-White), White with diabetes (Dx-White), African Americans without diabetes (NoDx-AA), and African Americans with diabetes (Dx-AA). Comparison groups were equally distributed in terms of sex of participants. Comparison groups were also matched for body mass index (BMI), age, and poverty status.

|  | NoDx-White<br>(N=10) | Dx-White<br>(N=10) | NoDx-AA<br>(N=10) | Dx-AA<br>(N=10) | Overall<br>(N=40) |
| --- | --- | --- | --- | --- | --- |
| **Sex** |  |  |  |  |  |
| - Women | 5 (50%) | 5 (50%) | 5 (50%) | 5 (50%) | 20 (50%) |
| - Men | 5 (50%) | 5 (50%) | 5 (50%) | 5 (50%) | 20 (50%) |
| **Body Mass Index (BMI)** |  |  |  |  |  |
| Mean (SD) | 29.46 (3.75) | 32.54 (5.99) | 30.03 (5.67) | 29.03 (3.58) | 30.27 (4.88) |
| Median | 30.47 | 33.86 | 27.64 | 29.35 | 29.76 |
| [Min, Max] | [22.66, 34.29] | [24.78, 41.28] | [23.18, 40.71] | [20.57, 34.01] | [20.57, 41.28] |
| **BMI category** |  |  |  |  |  |
| (20,30] | 4 (40%) | 4 (40%) | 6 (60%) | 6 (60%) | 20 (50%) |
| (30,42] | 6 (60%) | 6 (60%) | 4 (40%) | 4 (40%) | 20 (50%) |
| **Age** |  |  |  |  |  |
| Mean (SD) | 51.35 (11.31) | 49.53 (8.53) | 52.3 (11.13) | 54.56 (10.47) | 51.93 (10.18) |
| Median | 55.6 | 52.1 | 57.45 | 58.25 | 55.45 |
| [Min, Max] | [32.5,64.7] | [34.4,58.1] | [30, 61.9] | [37.4,64.8] | [30.0, 64.8] |
| **Poverty status** |  |  |  |  |  |
| Above | 9 (90%) | 7 (70%) | 9 (90%) | 7 (70%) | 32 (80%) |
| Below | 1 (10%) | 3 (30%) | 1 (10%) | 3 (30%) | 8 (20%) |

Shipment of plasma and peripheral blood mononuclear cells (PBMCs) samples used for this study was coordinated by HANDLS study coordinators from the National Institute on Aging, NIH, Baltimore to the University of California, Irvine (UCI). Cryopreserved plasma and PBMCs were stored upon arrival at the Nicholas laboratory at UCI in a -80C freezer and a liquid nitrogen (LN) tank, respectively. Plasma samples were thawed on ice before processing for targeted lipidomics and multiplex cytokine profiling. PBMCs were thawed in a water bath at 37°C prior to in vitro activation and staining for flow cytometry.

### AllofUS Population study and experimental design

In addition to using a HANDLS subcohort, we used information on clinical data related to lipids and inflammation from a T2D subcohort from the AllofUs multi-ethnic study (Ramirez et al., 2022). The AllofUs study is an NIH funded multicenter study consisting of patient-level data from 331,382 individuals from 35 hospitals in the United States aimed at sampling one million or more people living in the United States to provide a collection of broadly accessible data. The AllofUs research program reflects the diverse population in the United States, including minority groups historically underrepresented in biomedical research. We used the AllofUs researcher workbench to create a dataset of 17,339 participants with T2D from two different racial groups (White and AA) from the same age and body mass index ranges as our HANDLS subcohort (Registered Tier Data v.7). We further selected the AllofUs T2D subcohort based on disease status (T2D), race, age (range matched to HANDLS: 30 to 65 yo), and BMI (range matched to HANDLS: 20 to 42). We evaluated differences in glycated hemoglobin (HbA1C), insulin, glucose, cholesterol to HDL ratio (CholHDLRat), triglycerides, and C-Reactive protein (CRP) between White and African American cohorts.

The institutional review board of the National Institute of Environmental Health Sciences and the National Institutes of Health approved these protocols. The University of California Irvine Institutional Review Board exempted this study from review.

### Demographics and clinical parameters, HANDLS study cohort

Demographic and clinical parameters relevant to diabetes were provided to us by HANDLS study coordinators. The data provided was from a cross-sectional sample collected between 2004 and 2008. After informed consent and enrollment into the study, participants were interviewed and subjected to an examination on a mobile research vehicle. Height and weight were measured to calculate body mass index, whole blood was processed and stored as serum, plasma, and peripheral blood mononuclear cells (PBMCs). Disease status, race, sex, age, body mass index, and poverty status were used to equally distribute and match our comparison groups as follow: non-diabetic White (NoDx-White), diabetic White (Dx-White), non-diabetic African Americans (NoDx-AA), and diabetic African Americans (Dx-AA). Clinical measurements used for analysis were related to metabolic homeostasis and systemic inflammation and include cholesterol, high-density lipoprotein (HDL), cholesterol/HDL ratio, triglycerides, low-density lipoprotein, very-low-density lipoprotein, HbA1c, waist hip ratio, high sensitivity C-reactive protein (hs-CRP), insulin, and fasting blood glucose.

### Dietary recall data

Dietary intake of lipids was determined using the USDA Automated Multiple Pass Method of dietary recall, which is an interviewer-administered computerized method for collecting 24-hour dietary recalls (U.S. Department of Agriculture, n.d.)

### Clinical Data analysis

Univariate analysis on clinical measurements was performed using two-way ANOVA and post-ANOVA comparisons using Fisher’s LSD test. In addition, we performed principal component analysis (PCA) of clinical variables to assess factors contributing to variability of groups. PCA is a mathematical algorithm that determines which factors correlate with each other and which factors contribute the most and the least to variability within the dataset. The squared cosine (Cos2) is the parameter of a PCA that shows the importance of a component for any given observation (Ringnér, 2008).

### Targeted Lipidomics using Liquid Chromatography Mass Spectrometry (LC-MS)

#### Metabolite Extraction

To extract metabolites from plasma samples, 300μL -20°C 1000:1 isopropanol:lipidomics standard (extraction solvent) was added to 10μL of aliquoted plasma sample and incubated on ice for 10 min, followed by vortexing and centrifugation at 15,800 x g for 15 min at 4°C. 100μL of the clear supernatant (extract) was transferred to a glass mass spectrometry vial.

#### LC-MS

Plasma extracts were analyzed by LC-MS. Metabolites were analyzed using a quadruple-orbitrap mass spectrometer (Q-Exactive Plus Quadrupole-Orbitrap, Thermo Fisher) coupled to reverse-phase ion-pairing chromatography. The mass spectrometer was operated in positive ion mode with resolving power of 140,000 at m/z 200 and scan range of m/z 290-1200. The LC method utilized an Atlantis T3 column (150 mm x 2.1 mm, 3 μm particle size, 100Å pore size, Waters) with a gradient of solvent A (90:10 water: methanol with 1mM ammonium acetate and 35 mM acetic acid) and solvent B (98:2 Isopropanol: methanol with 1mM ammonium acetate and 35 mM acetic acid). The LC gradient was 0 min, 25% B, 0.150 ml/min; 2 min, 25% B, 0.15 ml/min; 5.5 min, 65% B, 0.150 ml/min; 12.5 min, 100% B, 0.150 ml/min: 16.5 min, 100% B, 0.150 ml/min, 17 min, 25% B, 0.150 ml/min and 30 min, 25% B, 0.150 ml/min. Other LC parameters were column temperature 45C, autosampler temperature was set to 4°C and the injection volume of the sample was 3μL.

#### Data analysis

Lipidomics data analysis was performed with Compound Discover and MAVEN software.

### Cytokine profiling using Luminex platform

#### Plasma samples

One aliquot per donor of frozen plasma samples from the HANDLS cohort (N=40) were thawed on ice to perform cytokine profiling using the Luminex Intelliflex platform. Plasma samples were used in a 1:100 dilution using serum matrix provided with MILLIPLEX^®^ MAP human kits used and in a neat state (no dilution) for comparison to assess background signal. 10µl of each sample (diluted and neat) were evaluated per kit in a 384-well plate for cytokine profiling.

#### Cytokine concentration measurements

For performance of cytokine profiling of plasma samples from the HANDLS cohort, we used the MILLIPLEX^®^ MAP human kits Cytokine/Chemokine/Growth Factor Panel A 47-plex (EGF, eotaxin, FGF-2, FLT-3L, fractalkine, G-CSF, GM-CSF, GROa, IFNa2, IFNy, IL-1a, IL-1b, IL-1ra, IL-2, IL-3, IL-4, IL-5, IL-6, IL-7, IL-8, IL-9, IL-10, IL-12p40, IL-12p70, IL-13, IL-15, IL-17a, IL-17e, IL-17f, IL-18, IL-22, IL-27, IP-10, MCP-1, MCP-3, M-CSF, MDC, MIG, MIP-1a, MIP-1b, PDGF-AA, PDGF-AABB, sCD40L, TGFa, TNFa, TNFb, VEGF-A) + 1 customized (RANTES) (Millipore Cat# HCYTA-60K-PXBK48) and the Th17 5-plex (customized: IL21, IL23, IL31, IL33, and MIP-3a) (Millipore Cat# HTH17MAG-14K) total 53 cytokines assayed per sample. To adjust the manufacturer’s protocol to our 384-well plate format all other reagents including antibodies, magnetic beads, and detection reagents, were used at 10 µL. Outcomes from wells with <50 beads for each analyte were excluded from analysis. Plates were washed in between incubations using a Biotek 450S touch plate washer (Biotek) and read using a xMAP INTELLIFLEX^®^ System (Luminex).

#### Data analysis

Quality control was performed on cytokine quantification data using the Belysa^®^ Immunoassay Curve Fitting Software (Millipore). This consisted of evaluating standard curves for all 53 analytes and quality control samples (provided in each kit) measured in the experiment. The interpolated concentrations of the plasma samples were exported in an excel file format for statistical and bioinformatic analysis (univariate, multivariate: K-means and gap stats, PCA, and OPLS-DA).

### Immune phenotyping of cellular populations using Flow cytometry

#### Tissue Culture

PBMCs were quickly thawed in a 37°C water bath and washed with 10mL of RPMI 1640 media (Genesee Scientific, CAT#25-506) supplemented with 10% Fetal Bovine Serum (Omega Scientific, CAT#FB-1), 20µM HEPES (ThermoFisher Scientific, CAT# 15630080), and 100 U/mL penicillin and 100µg/mL streptomycin (Genesee, CAT#25-512). Individual donor samples were split in half, one half was used for stimulation and the half left was used for immediate staining and flow cytometry acquisition. Half of the PBMCs for stimulation purposes were seeded at 1×10^6^ in a 96-well U-bottom plate coated with anti-human CD3/28 (Thermo Scientific CAT#501129356 & 501129714). Each donor PBMC sample (N=40) were plated in technical duplicates. CD3/28 activated PBMCs were cultured for 40 hours in 37°C 5% CO_2_ followed by sample preparation and analysis by flow cytometry.

#### Flow cytometry

A panel of 22 surface and intracellular markers and 1 viability dye were used to phenotype various leukocyte populations in PBMCs (**Table 2**). PBMCs were first stained with live/dead stain Zombie NIR diluted in PBS, pH 7.4 for 20 minutes at 4°C away from light. PBMCs were washed with FACS buffer (PBS + 0.1% BSA + 2µM EDTA) and centrifuged at 500g for 5 minutes. Supernatant was removed and 25µL of Human TruStain FcX was added for 10 minutes on ice and protected from light to block human FC receptor to prevent nonspecific antibody staining. After 10 minutes, 25µL of surface antibody master mix diluted in BD Biosciences Brilliant Stain Buffer (CAT# 566349) was added to PBMCs for 20 minutes on ice away from light. PBMCs were then washed with FACs buffer, centrifuged and supernatant removed. PBMCs were then fixed with Biolegend’s Fixation Buffer (CAT# 420801) for 20 minutes at 4°C and protected from light. PBMCs were washed with Biolegend’s Intracellular Staining Permeabilization Wash Buffer (CAT# 421002) twice and supernatant was removed for the addition of 25µL of intracellular antibody master mix diluted in Biolegend’s Intracellular Staining Permeabilization Wash Buffer for 20 minutes at 4°C and protected from light. PBMCs were washed with Biolegend’s Intracellular Staining Permeabilization Wash Buffer and resuspended in 200uL of 1% paraformaldehyde diluted in PBS pH 7.4 for acquisition using the spectral flow cytometer Cytek’s 3-laser Northern Lights.

**Table 2.** Flow cytometry panel used to phenotype cellular populations in PBMCs.

| Marker | Fluorophore | Clone | Purpose | Supplier | Cat. # | Dilution |
| --- | --- | --- | --- | --- | --- | --- |
| CCR7/<br>CD197 | PE/Fire 810 | G043H7 | Memory T cells | Biolegend | 353269 | 200 |
| CD11c | Pacific Blue | 3.9 | Dendritic cells | Biolegend | 301626 | 100 |
| CD14 | PE/Cyanine7 | 63D3 | Monocytes | Biolegend | 367112 | 100 |
| CD16 | BUV 805 | 3G8 | NKC & monocyte | BD<br>Biosciences | 748850 | 400 |
| CD19 | AF647 | HIB19 | B cells | Biolegend | 302220 | 400 |
| CD25 | APC-R700 | 2A3 | Regulatory T<br>cells | BD<br>Biosciences | 565106 | 200 |
| CD3 | PerCP | OKT3 | T cells | Biolegend | 317338 | 200 |
| CD4 | Spark YG 593 | SK3 | helper T cells | Biolegend | 344672 | 100 |
| CD45 | BV 510 | 2D1 | Pan leukocytes | Biolegend | 368526 | 400 |
| CD45RA | BV 785 | HI100 | Naive T cells | Biolegend | 304140 | 400 |
| CD56 | BUV496 | NCAM16.2 | NK cells | BD<br>Biosciences | 750479 | 50 |
| CD69 | BUV 563 | FN50 | T cell activation | BD<br>Biosciences | 748764 | 100 |
| CD8 | Spark Blue 550 | SK1 | Cytotoxic T cells | Biolegend | 344760 | 200 |
| FOXP3 | PE-Cy5.5 | PCH101 | Treg | ThermoFish<br>er Scientific | 35-4776-42 | 50 |
| HLA-DR | BV711 | L243 | Antigen<br>presenting cells | Biolegend | 307644 | 100 |
| PD1 | Super Bright<br>645 | MIH4 | T cell activation | ThermoFish<br>er Scientific | 64-9969-42 | 100 |
| TIGIT | BUV615 | 741182 | T cell exhaustion | BD<br>Biosciences | 752314 | 100 |
| IgM | BUV395 | G20-127 | B cell | BD<br>Biosciences | 563903 | 400 |
| IgD | BUV661 | IA6-2 | Mature B cells | BD<br>Biosciences | 741637 | 400 |
| CD45RO | BUV737 | UCHL1 | Memory T cells | BD<br>Biosciences | 748368 | 100 |
| CD62L | APC-Fire 810 | DREG-56 | T cell subsets | Biolegend | 304866 | 400 |
| CD44 | Nova Fluor Blue<br>610/70S | IM7 | T cells | ThermoFish<br>er Scientific | M010T02<br>B06 | 100 |
| Live/Dead | Zombie NIR | --- | Cell viability | Biolegend | 423106 | 3200 |
| Fc Receptor | Human TruStain<br>FcX | --- | Block human FC<br>receptors to<br>prevent false<br>positives or false<br>negatives | Biolegend | 422302 | 50 |

**Table 3.**
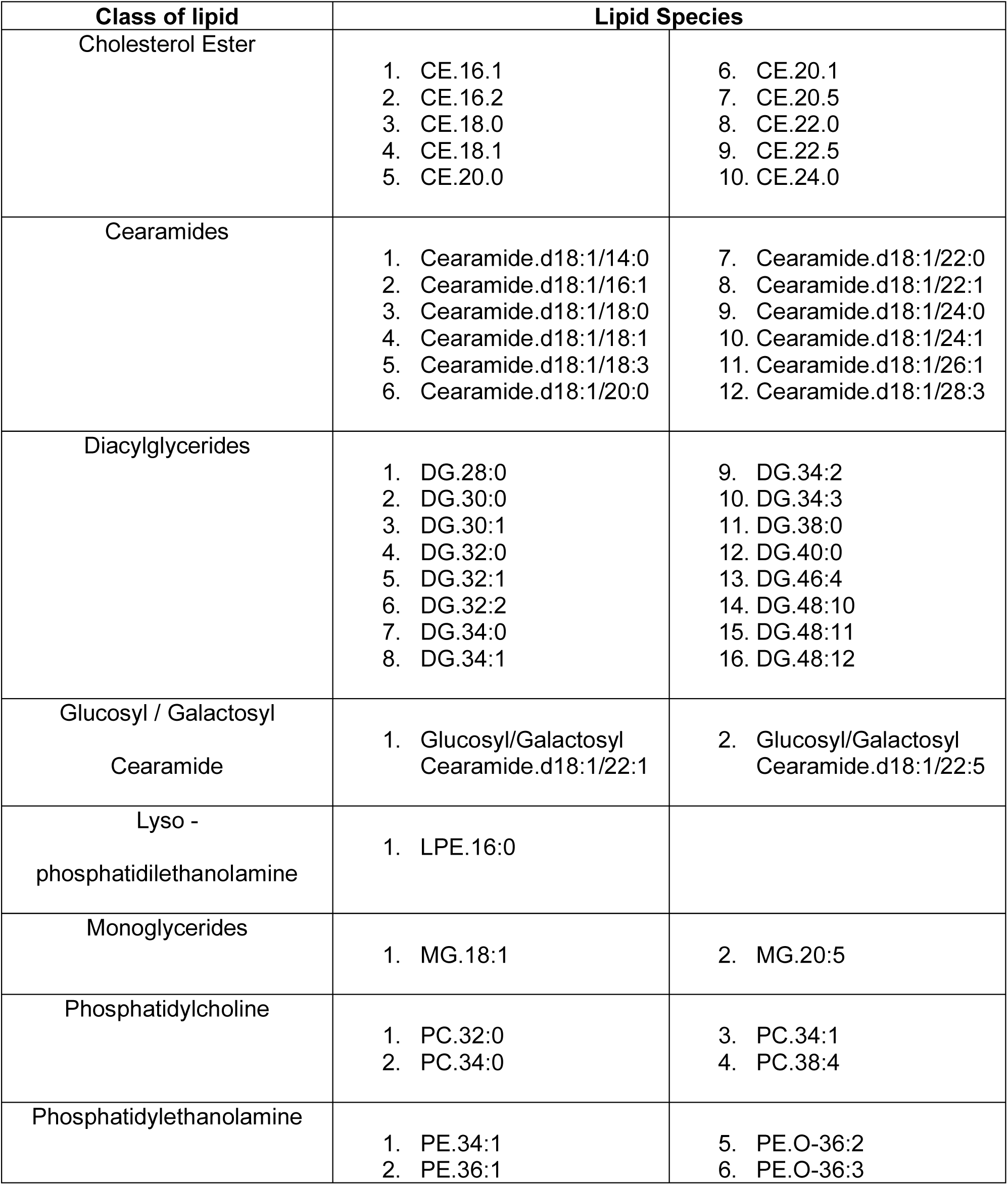
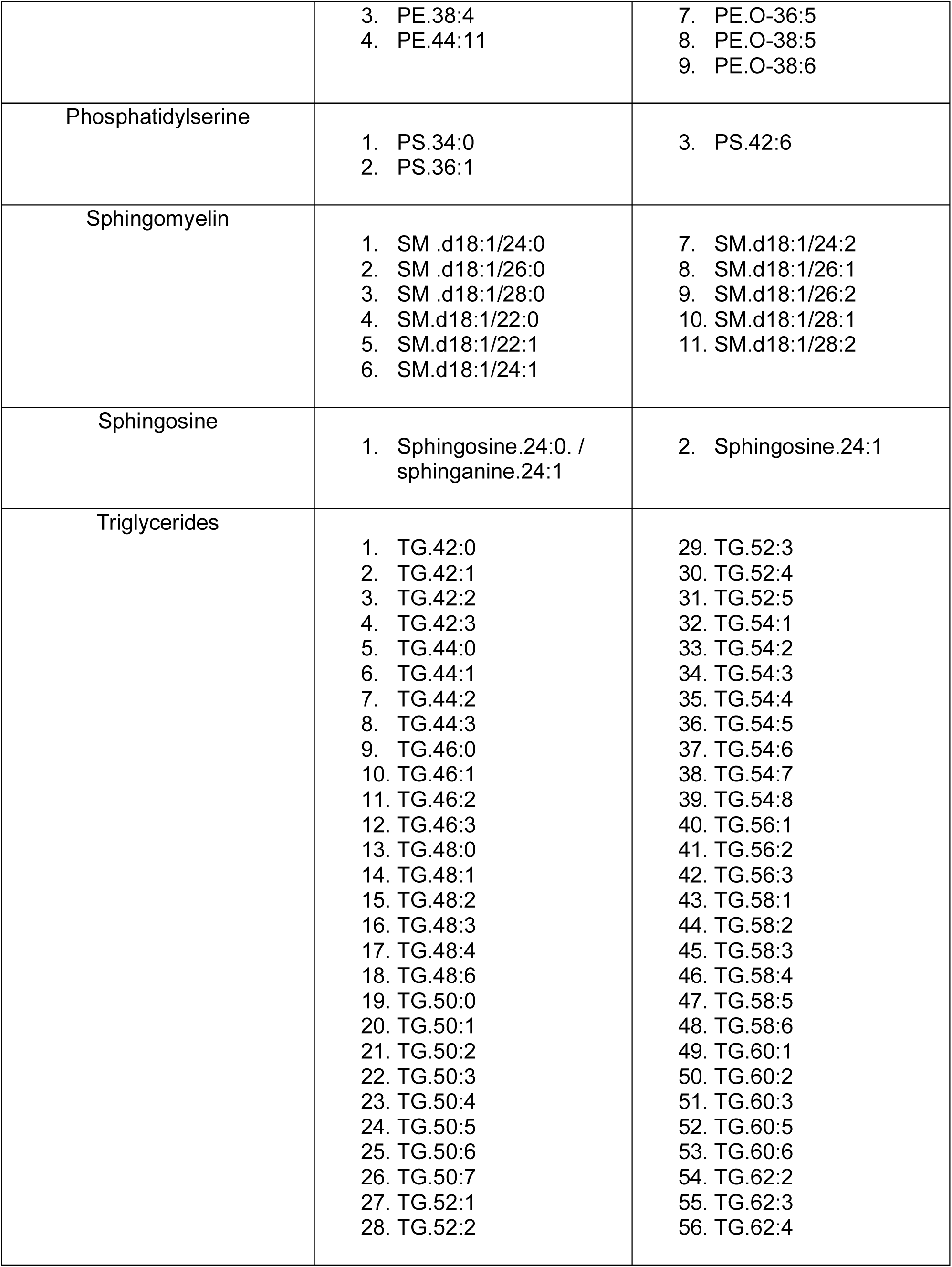
List of 128 lipids evaluated using targeted lipidomics in HANDLS subcohort. **Table legend:** A subcohort of 40 individuals from the HANDLS study were divided into 4 comparison groups based on disease status and race: White without diabetes (NoDx-White), White with diabetes (Dx-White), African Americans without diabetes (NoDx-AA), and African Americans with diabetes (Dx-AA).

#### Data analysis

Flow Cytometry data was analyzed with FlowJo v.10. Cells were gated first by time of sample acquisition followed by doublet discrimination using FCS-H and FSC-A. Next live cells were gated based on viability dye, then for size and granularity for lymphocytes and myeloid cells based on SSC-A and FSC-A. Boolean gating was used for each population of interest based on their specific markers, for example CD3+CD4+ for CD4+ T cell, CD3+CD8+ for CD8+T cells, etc. Percentages of cell populations (frequencies) from the parent population and from the total of live immune cells were exported from the software and uploaded in R Studio and GraphPad prism v.10 for statistical analysis and figure generation.

### Statistical analysis ANOVA model in HANDLS subcohort

Statistical analysis, specifically two-way ANOVA and post-ANOVA multiple comparisons were performed using R studio and GraphPad prism. Briefly, all datasets (clinical parameters, targeted lipidomics, and plasma cytokines) were assessed for normal distribution. Then, for targeted lipidomics and plasma cytokines, we performed a box-cox transformation on the datasets as needed to correct for heteroscedasticity and to better satisfy the normality assumption of the two-way ANOVA model. We then fit the model considering disease status and race as our factors along with the interaction between the two and evaluated the main effects and interaction effects through the F-test on the sums of squares. After determining which measurements were significantly different in an overall model considering disease status, race, and the interaction between the two, we performed post hoc multiple comparison analysis using Fisher Least Significant Difference (LSD) test and plotted the results. All graphics were generated using GraphPad Prism v. 10.

### Feature selection analysis using Orthogonalized Partial Least Squares discriminant analysis (OPLS-DA)

OPLS-DA is an iteration of the supervised clustering approach partial least squares discriminant analysis (PLS-DA), similar to the more conventionally used principal component analysis (PCA). OPLS-DA is a machine learning tool that selects features characteristic in a training dataset to build a predictive model tested on a validation dataset (Tapp & Kemsley, 2009; Trygg & Wold, 2002; Worley & Powers, 2012). OPLS-DA generates latent variables (LVs), like PLS-DA, that are analogous to the principal components obtained by PCA but constrained by categorical information. Different from PLS-DA, OPLS-DA applies orthogonal rotations to the analysis to obtain maximum separation of classes along the LV1 axis, hence a single LV serves as a predictor for the class, while other components describe the variation orthogonal to the first predictive component (LV1).

#### Analysis

OPLS-DA was performed for both datasets independently (targeted lipidomics and cytokine profiling) using the Solo eigenvector research software. Due to the sample size of our HANDLS subcohort (40 participants), we limited the application of this tool to build a feature selection model using our dataset as a calibration set only, an approach conducted in previously published work (Barroeta-Espar et al., 2019; Wood et al., 2015). This allowed for the discovery of distinct immunometabolic features of diabetes between White and African American cohorts. Each dataset was normalized (z-scored) before uploaded to the Solo software. Classes selected for comparison were based on disease status (with and without diagnosis of diabetes) in each racial group (African American and White). For example, diabetes group comparing classes as White vs African American cohorts. Cross-validation was performed using the leave-one-out cross-validation strategy, available in the Solo software. Performance of the feature selection model generated was evaluated by statistics R^2^ Cal or R^2^ calibration which refers to the coefficient of determination for the calibration set and measures the proportion of variance in the response variable that can be explained by the model using the calibration set. A higher R^2^ Cal value indicates a better fit of the model for the calibration data, suggesting that the model captures the underlying patterns well. LV1 and LV2 loadings were used to plot the datasets and orthogonalized LV1 loadings were used to determine features that correlated positively and negatively with disease status in each racial group.

### Correlative analysis of lipid:cytokine ratios to clinical markers of diabetes

The ratio of every possible lipid:cytokine combination was computed for all subjects of the HANDLS subcohort (N=40). The ratios and their significance were generated using the corAndPvalue() function from the WGCNA package (version 1.72-5) in R studio (version 4.2.1). Further, we evaluated which lipid:cytokine ratios uniquely correlated to HbA1c, a clinical parameter used to diagnose diabetes, and HOMA-IR, a measurement of insulin sensitivity, in each group. We filtered for lipid:cytokine ratios that correlated significantly (p-value < 0.05) with HbA1C and HOMA-IR in at least one racial group. We plotted correlation statistics for these ratios each for White and African American individuals using ggplot2 (version 3.5.1).

### Demographics and health data from the AllofUs study cohort

#### Data Extraction

Data for this study were extracted from the AllofUs study using the AllofUs Researcher workbench platform. The dataset was queried focusing on specific demographic and health information. Custom SQL queries were used to extract data related to measurement concepts of interest (e.g., biomarker levels), condition occurrences, and participant demographic details such as race and sex. The data were exported to the online platform Rstudio, available in the AllofUs Researcher Workbench, allowing for efficient access and analysis.

#### Data analysis

Once imported, the data underwent preliminary cleaning, such as removal of missing values and unnecessary columns. After this, we computed descriptive statistics, including the mean, standard deviation, and standard error of the mean (SEM) for each clinical biomarker stratified by racial group. These statistics were presented in a tabulated format using the kable package. To adjust biomarker levels for potential confounders (body mass index and age) across different racial groups, a custom R function was implemented. This function fitted linear models to each biomarker of interest, adjusting for BMI and age while accounting for race. Adjusted biomarker levels were calculated by subtracting the contributions of BMI and age, as determined by the linear model coefficients. Subsequent t-tests were performed to compare the adjusted biomarker levels between racial groups, excluding the racial group Asian due to insufficient data values. The results were visualized using ggplot2 and ggpubr, with t-test statistics annotated on box plots that displayed the adjusted biomarker levels by race. All analyses were conducted in R, with statistical significance set at p < 0.05.

## RESULTS

### Overall study design

We generated three independent biological datasets for integrated analysis from the HANDLS subcohort (**Fig. 1**). First, we analyzed serum clinical parameters (**Fig. 1A and 1B**). Second, we used plasma samples from HANDLS study participants to perform targeted lipidomics using liquid chromatography mass spectrometry (LC-MS) and to perform cytokine and growth factor multiplex quantification using Luminex. These approaches resulted in 128 lipids and 47 cytokines for analysis (**Fig. 1C and 1D**). Within each data type, we performed univariate and multivariate analysis among our 4 study groups. Finally, we performed an integrated analysis of all biological datasets to identify clinical and immunometabolic features characteristic of diabetes in either White or African American populations (**Fig. 1E**).

**Figure 1.**
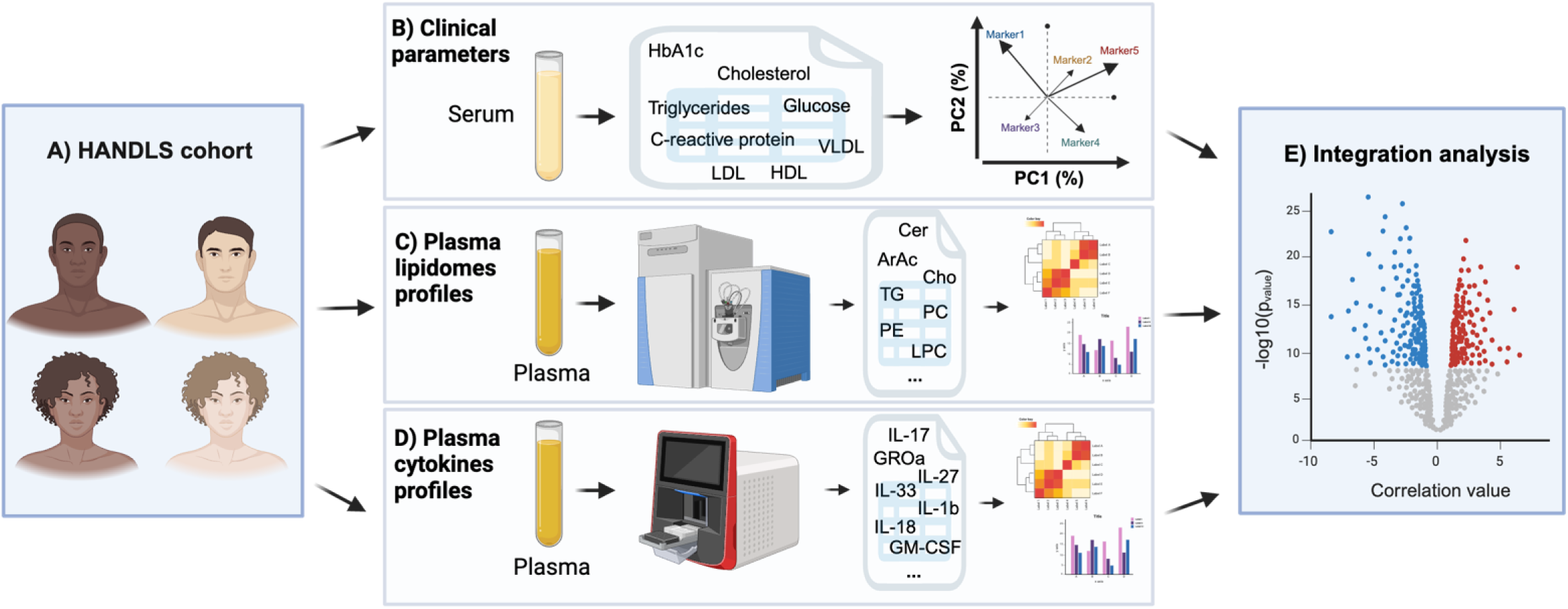
Study experimental design using a diverse HANDLS subcohort. A) HANDLS cohort schematic showing equal distribution of participants by race and sex. Participants were also equally divided by diabetes status. Techniques employed for the generation of datasets are shown in B, C, and D. B) Clinical parameters measured in serum consisted of glucose measurements (HbA1C and glucose), lipids measurements (cholesterol, triglycerides, HDL, LDL, and VLDL), and inflammation measurements (C-reactive protein). C) Plasma lipidomes profiles were generated using targeted lipidomics. D) Plasma cytokines profiles were generated using multiplex Luminex platform. All independent analysis consisted of statistical and bioinformatic assessment and visualization tools. Lastly, E) Integrative analysis of all datasets generated was performed.

### Clinical lipids are major drivers of variability in the HANDLS subcohort

To define the clinical features characteristic of diabetes in White and African American participants, we first performed univariate comparison of clinical parameters. We evaluated waist hip ratio (WHR), cholesterol (Chol) levels, high-density lipoprotein (HDL), cholesterol to HDL ratio (CholHDLRat), low-density lipoprotein (LDL), very low-density lipoprotein (VLDL), triglycerides, HbA1C, insulin, fasting glucose, and high sensitivity C-reactive protein (hs-CRP). We performed a two-way ANOVA to determine whether disease (diabetes status), race, and/or the interaction of both variables modify the clinical parameters in our cohort (**Fig. 2A**). We found that CholHDLRat, VLDL, HbA1C, hs-CRP, insulin, and glucose levels were significantly modulated by diabetes status. Insulin was the only parameter that was significantly modulated by race in our statistical model (**Fig. 2A**). Though insulin was not significantly different in any individual comparisons (**Suppl. Fig S1A**), insulin levels were significantly different between NoDx-White and Dx-White when adjusted for insulin use (**Suppl. Fig S1B)**. Interestingly, hs-CRP was the only parameter that was significantly modulated by the interaction of disease and race. Confirming our statistical model (**Fig. 2A**), standard-of-care clinical measurements for diagnosis of diabetes such as HbA1C and fasting glucose, were significantly different between individuals with and without diabetes in each racial group independently (**Fig. 2B**).

**Figure 2.**
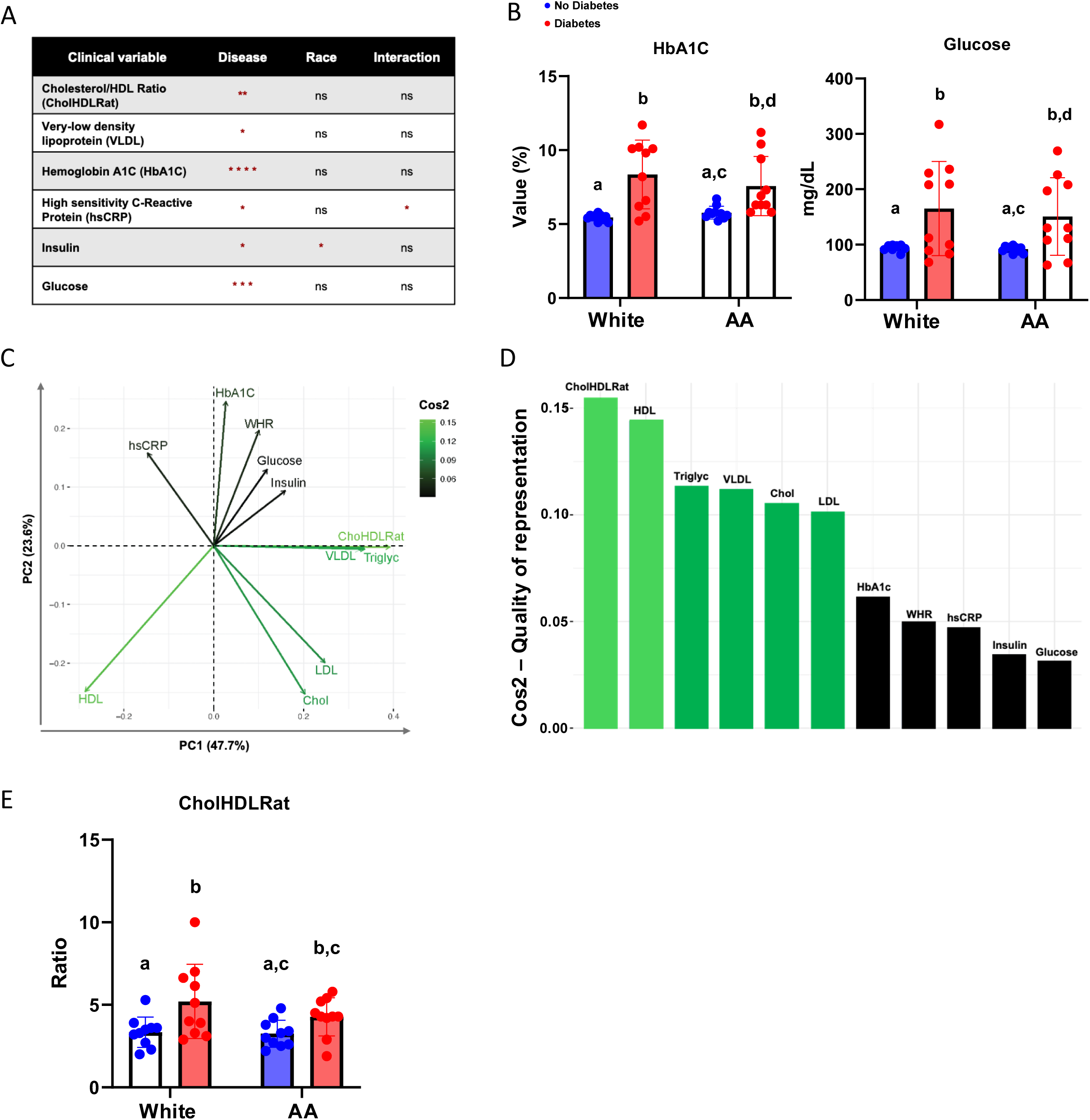
Clinical lipids are main drivers of variability in a diverse HANDLS subcohort. A) Table showing clinical parameters that differed statistically based on disease (no diabetes vs diabetes), race (White vs AA), and on the interaction of both variables (ns= not significant, * = p-value<0.05, ** = p-value<0.01, *** = p-value<0.001, **** = p-value<0.0001). B) Bar/dot graph showing results from multiple post-anova comparison of HbA1C (left) and fasting glucose (right) among white and AA with and without diabetes. C) Principal Component Analysis (PCA) showing correlations among clinical parameters evaluated and Cos2 color gradi ent indicating the quality of representation of clinical parameters of PCA from lowest to highest (black to lightest green). D) Contribution bar chart displaying the order of parameters contributing to variability from highest to lowest (highest light green bar to lowest black bar) based on Cos2. E) Bar/dot graph showing results from multiple statistical comparison of Cholesterol/HDL ratio (CholHDLRat) among White and AA with and without diabetes. Blue = People without diabetes, red = people with diabetes. Statistical analysis performed using Two-way ANOVA with Box Cox transformed values followed by Fisher’s LSD post-comparison test (unadjusted p-values). Statistical post-hoc comparisons were performed only between matched groups based on diabetes status and race and comparisons between persons with and without diabetes were not included in the analysis. P-values obtained from multiple post-hoc comparison analysis are represented using a statistical letter system, where significantly different p-values are represented by different letters and non-significant p-values are represented by the same letters.

To better understand the sources of variation in the clinical parameters evaluated in our diverse HANDLS subcohort, we performed principal component analysis (PCA) (**Fig. 2C and 2D**). PCA is a mathematical algorithm that determines which factors contribute the most and least to variability within the dataset (Ringnér, 2008). By projecting the variables on principal component 1 (PC1), we determined that clinical lipid measurements like CholHDLRat, HDL, and triglycerides were the top 3 contributors to variability in our cohort (**Fig. 2D**). Counterintuitively, standard-of-care diabetes measurements such as HbA1C, fasting glucose, and insulin contributed the least to the variability evaluated in our cohort. Intriguingly, CholHDLRat, the main driver of variability, was only significantly different when compared between individuals with and without diabetes in the White group, even after adjusting for lipid-lowering drug use (statins) which could affect cholesterol levels (**Fig. 2E** and **Suppl Fig. S2**). From these data, we conclude that variability in diabetes associated with clinical parameters is lipid driven.

### Plasma lipidomes characterize diabetes in White but not in African American groups in the HANDLS subcohort

Intrigued by the results indicating that lipid measurements are main drivers of variability in the HANDLS subcohort (**Fig. 2C and 2D**) and that CholHDLRat distinguishes diabetes from non-diabetes status in White but not in African American cohorts (**Fig. 2E)**, we performed a more in-depth analysis of dietary and endogenous lipids in the HANDLS subcohort. First, we evaluated the dietary intake of lipids, as measured by the USDA Automated Multiple Pass Method of dietary recall. No differences were observed when comparing intake of lipids: fats (mono, poly, and saturated) and fatty acids (short, medium, and long chain fatty acids) between Dx-White and Dx-AA and in each group. We only observed slight differences when comparing some short-chain fatty acids between NoDx-White and NoDx-AA (**Suppl. Table S1**). Next, we performed targeted plasma lipidomics to identify specific endogenous lipid species that could be differentially abundant across our comparison groups (NoDx-White, Dx-White, NoDx-AA, and Dx-AA). Similar to our analysis of clinical measurements (**Fig. 2A**), we performed two-way ANOVA to assess whether disease (diabetes status), race, and/or both variables significantly modulated the differences in lipid abundance seen in our HANDLS subcohort. We found that out of 128 lipids measured through targeted plasma lipidomics, 38 lipids were significantly modulated in a model where disease, race, or both variables were evaluated (**Fig. 3A**). Majority of these lipids were triglycerides (TG) (**Fig. 3B**). We continued performing multivariate analysis using K-means and gap statistics on the significantly modulated lipids. By comparing cluster centers, a measurement that represents the average expression of all correlated lipids in one cluster, we found that all 3 clusters generated were significantly different in at least one comparison performed (**Fig. 3B**). Cluster 1, comprising mainly polyunsaturated long chain triglycerides (TG) phosphatidylcholine (PC), phosphatidylethanolamine (PE), and sphingosine, was increased in White individuals over the African American group regardless of diabetes status (**Fig. 3C**). Cluster 2 was increased in Dx-AA and Dx-White compared to NoDx-AA and NoDx-White, respectively (**Fig. 3D**). Cluster 3, composed of mainly long chain diacylglycerides (DG), and very long chain TG, was increased in Dx-White compared to Dx-AA (**Fig. 3E**). We conclude from the targeted plasma lipidomics analysis that long and very long chain DG and TG are most abundant in and most impacted by diabetes status in the White participants.

**Figure 3.**
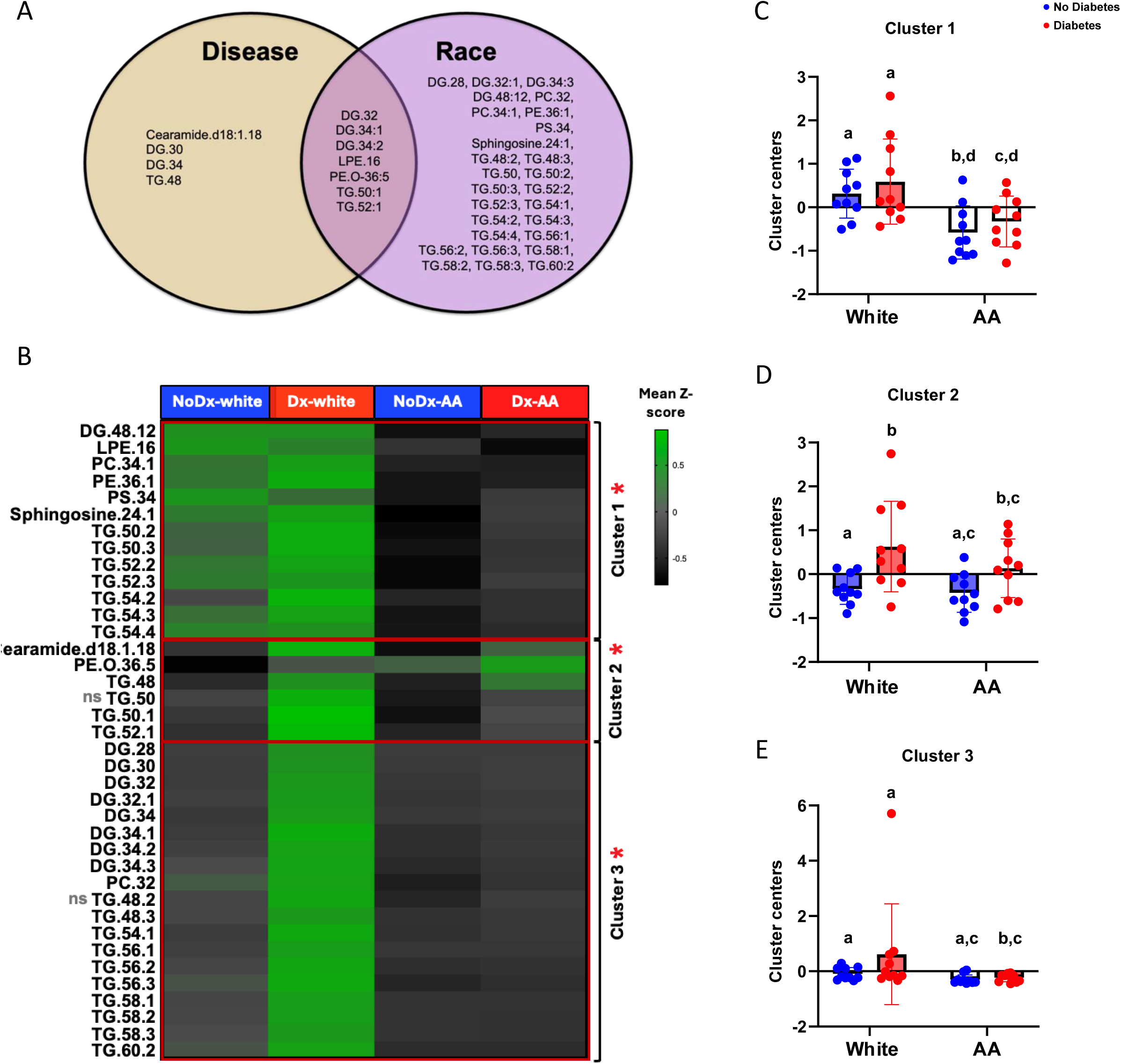
Plasma lipidomes characterize diabetes in White groups but not in AA groups in a diverse HANDLS subcohort. **A)** Venn diagram showing 38 lipids that were significantly modulated based on disease (no diabetes vs diabetes), race (White vs AA), and on both disease and race. **B)** Heatmap showing mean z-score value per comparison group from lowest (dark grey) to highest (light green) lipid species evaluated univariately and through cluster analysis (clusters 1, 2, and 3) generated using K-means and gap statistics. “ns” next to TG.50 and TG.48.2 represent non-significance in post-anova comparisons and red asterisks next to clusters represent statistically significant clusters. **C)** Bar/dot graph showing results from post-anova multiple comparison of lipid cluster 1 among White and AA with and without diabetes. **D)** Bar/dot graph showing results from post-anova multiple comparison of lipid cluster 2 among white and AA with and without diabetes. **E)** Bar/dot graph showing results from post-hoc multiple comparison of lipid cluster 3 among White and AA groups with and without diabetes. X axis represents cluster center measurements. Blue = People without diabetes, Red = people with diabetes. Statistical analysis performed using Two-way ANOVA with Box Cox transformed values followed by Fisher’s LSD post-comparison test (unadjusted p-values). Statistical post-anova comparisons were performed only between matching groups based on diabetes status and race and comparisons between persons with and without diabetes were not included in the analysis. P-values obtained from multiple post-hoc comparison analysis were represented using a statistical letter system, where significantly different p-values are represented by different letters and non-significant p-values are represented by the same letters.

### Classical measures of inflammation characterize diabetes in White but not in African American groups in HANDLS subcohort

From the lipid analysis, it became evident that the contribution of dyslipidemia to the presentation of diabetes in the African American cohort was different than in the White cohort. Because diabetes is not only a metabolic condition, but also an inflammatory disease (Calle & Fernandez, 2012), we decided to address whether diabetes-related inflammation was also manifesting differently in our populations. We were puzzled to see that hs-CRP, the most common clinical inflammatory marker (Blake & Ridker, 2002; Kalaiselvan et al., 2023; Lee et al., 2024), was not a significant contributor to variability in our dataset (**Fig. 2C and 2D**). Further, we were surprised to see that despite hs-CRP being significantly modulated in a model in which diabetes status and race were interactive variables (**Fig. 2A**), multiple comparison analysis showed that hs-CRP was only elevated in diabetes in the White group, even after adjusting for statin use which could alter hs-CRP levels **(Fig. 4A, Supp. Fig. S3**). These data prompted us to explore other systemic inflammatory biomarkers that could better characterize inflammation in this African American cohort and potentially reveal novel features of diabetes presentation in this population.

**Figure 4.**
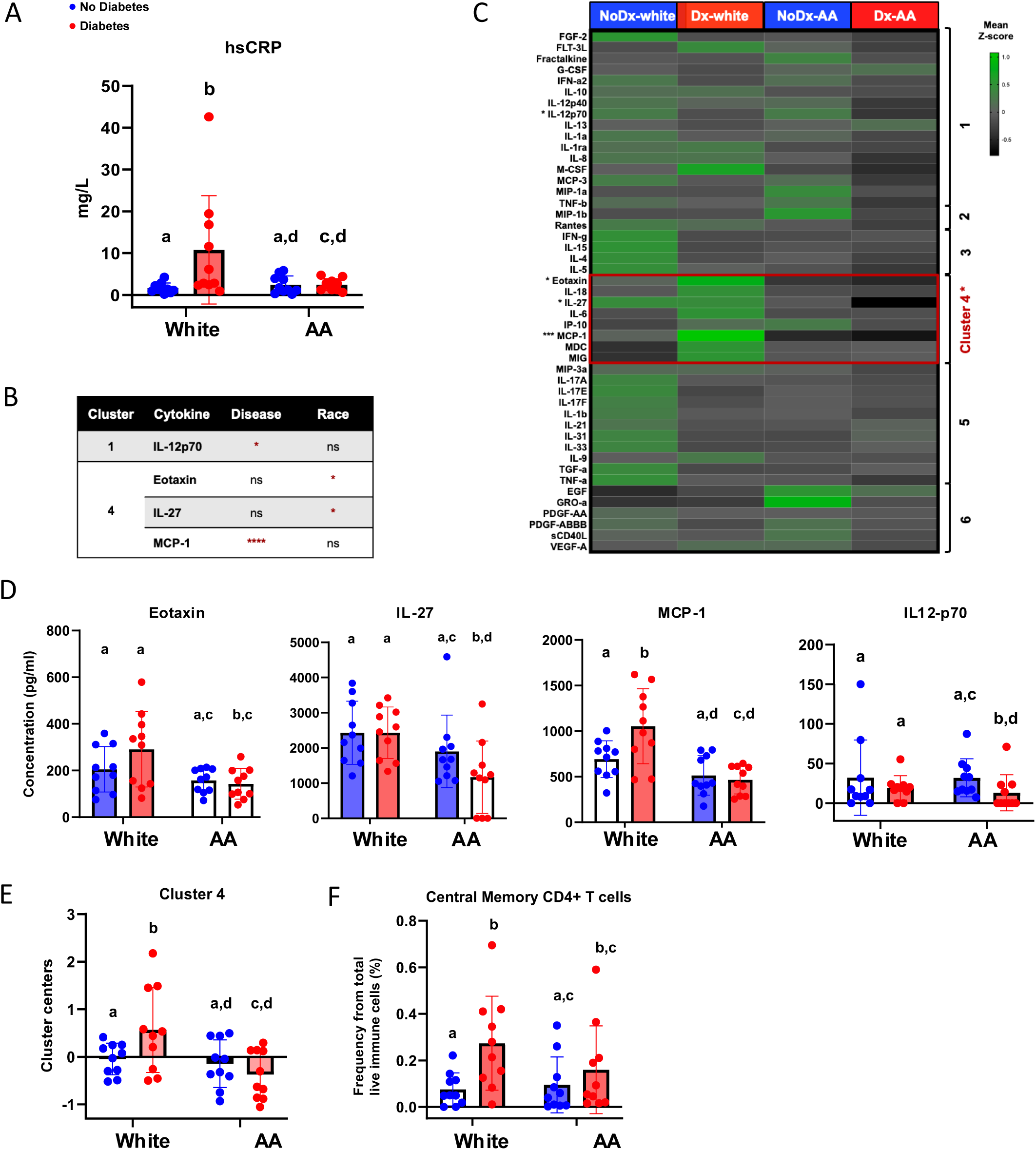
Plasma inflammatory profiles characterize diabetes in White but not in AA groups in a diverse HANDLS subcohort. **A)** Bar/dot graph showing results from multiple statistical comparison of high sensitivity C-reactive protein (hsCRP) among White and AA with and without diabetes. **B)** Table showing cytokines and their respective cluster that differed statistically based on disease (no diabetes vs diabetes), race (White vs AA) or interaction of the variables (ns= not significant, * = p-value<0.05, **** = p-value<0.0001). **C)** Heatmap displaying mean z-score value per comparison group of plasma cytokines evaluated and per cytokine cluster (generated by K-means and Gap statistics analysis). Asterisks next to cytokines and cluster of cytokines represent significantly modulated clusters. **D)** Bar/dot graph showing cytokines eotaxin (left), IL-27 (center), MCP-1, and IL12p-70 (right) which were statistically different among comparison groups. **E)** Bar/dot graph showing results from statistical comparison of cytokine cluster 4 among White and AA with and without diabetes. **F)** Bar/dot graph showing results from statistical comparison of cellular population central memory CD4+ T cells among White and AA with and without diabetes. X axis represents cluster center measurements. Blue = people without diabetes, Red = people with diabetes. Statistical analysis performed using Two-way ANOVA with Box Cox transformed values followed by Fisher’s LSD post-comparison test (unadjusted p-values). Statistical post-anova comparisons were performed only between matched groups based on diabetes status and race. Comparisons between persons with and without diabetes were not included in the analysis. P-values obtained from multiple post-anova comparison analysis were represented using a statistical letter system, where significantly different p-values are represented by different letters and non-significant p-values are represented by the same letters.

To determine whether additional systemic inflammatory proteins could be better discriminators of diabetes in African Americans, we performed multiplex cytokine and growth factor profiling using the Luminex platform. We probed for 52 analytes in plasma samples from our HANDLS subcohort (40 individuals). After quality control, we obtained concentration values for 47 molecules. Our initial statistical model assessment and univariate multiple comparisons among the 4 groups indicated that two cytokines were significantly modulated by disease status, IL-12p70 and MCP-1, and two other cytokines by race, eotaxin and IL-27 (**Fig. 4B and 4C**). Notably, all 4 cytokines (eotaxin, IL-27, MCP-1, and IL12-p70) were increased in Dx-White when compared to Dx-AA (**Fig. 4D**). Additionally, IL-27 and IL12-p70 were decreased in Dx-AA when compared to NoDx-AA and MCP-1 was increased in Dx-White compared to NoDx-White (**Fig. 4D**). Because cytokine production typically has a high probability of covariance, we performed a multivariate clustering analysis. Using K-means and gap statistics we generated 6 clusters representative of the 47 evaluated molecules. By comparing cluster centers, we determined that cluster 4, which included eotaxin, IL-27, and MCP-1, was significantly increased in Dx-White compared to all other groups (**Fig. 4C and 4E**). We conclude that immune cell function, as measured by systemic levels of specific cytokines eotaxin, IL-27, and MCP-1, could account for differences in diabetes inflammatory status among diverse populations.

Intrigued by the lack of a classical diabetes-associated inflammatory profile (i.e. IL-6, TNF-α, IL-1b, and hs-CRP) in the plasma of the African American cohort (**Fig. 4A and Suppl. Fig. S4**), we analyzed immune cell populations to determine if specific immune cell types could be contributing to this difference in inflammatory responses. We performed flow cytometry using a panel of 23 markers to phenotype immune cell populations and compare their frequencies among our 4 studied groups. Interestingly, central memory CD4^+^ T cells, a population of immune cells reported to play a modulatory role in diabetes (Rattik et al., 2019; Tan et al., 2022), was significantly increased only in Dx-White and not in Dx-AA (**Fig. 4F, Supp. Fig. S5**). No other phenotyped cell populations in human peripheral blood mononuclear cells (innate and adaptive cells) were significantly modulated amongst our 4 groups. We conclude that in our HANDLS subcohort, immune cell populations reported in literature to have changes in frequencies in diabetes could only characterize disease in White and not in the African American cohorts.

### Elevated lipids and classical inflammatory markers are features of diabetes in the White group while Th17 inflammatory features characterize diabetes in the African American group in the HANDLS subcohort

Conducting 4-group comparisons revealed lipids and inflammatory mediators were modulated distinctively in Dx-White. However, this approach did not allow us to detect markers of inflammation specific to Dx-AA. Therefore, we decided to use a supervised clustering approach to identify lipids and inflammatory features that characterize diabetes in both White and African American cohorts, and between individuals with and without diabetes of each racial group. We performed orthogonalized partial least squares discriminant analysis (OPLS-DA) to determine such features characterizing each racial group.

By performing feature selection with OPLS-DA using the set of 38 significantly modulated lipids from our targeted lipidomics diabetes diagnosed dataset, we first noticed a clear separation on latent variable 1 (LV1) driven by lipid profiles that correlated with disease status in African American or White groups (though the classification error for the cross-validation model was 0.5) (**Fig. 5A**). This separation between classes was even less clear when classifying racial groups independently by diabetes status (**Suppl. Fig S6A and S6C**). By comparing Dx-White from Dx-AA, we observed that the top 10 lipids that correlated positively with the presentation of diabetes in White individuals were long and very long chain TG in addition to monounsaturated species of phospholipids (**Fig. 5B**). Despite the classification model having high error, the lipid species identified using this model were significantly increased in Dx-White in comparison to NoDx-White and to Dx-AA (**Fig. 3B**). Additionally, some of the lipids reported to be markers of dyslipidemia, like ceramides and sphingosine, correlated positively with diabetes in White (both lipids) and African American (ceramides only) individuals (**Suppl. Fig S5B and S5D**). Our findings suggest that lipid profiles characterize the presentation of diabetes in White individuals, but not in African American individuals in this HANDLS subcohort.

**Figure 5.**
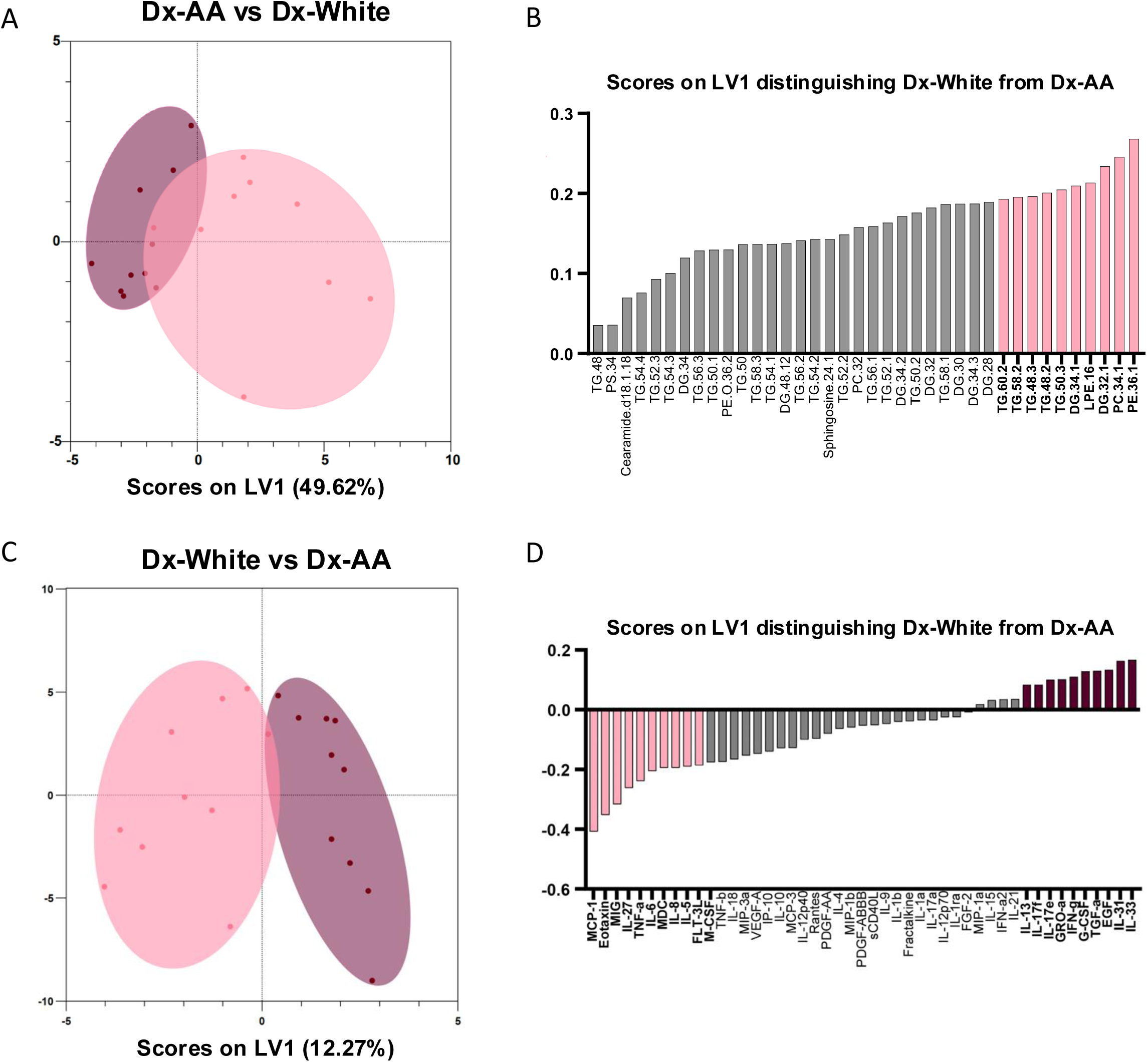
Lipid and inflammatory features characteristic of diabetes in a diverse HANDLS subcohort. **A)** Orthogonal partial least squares discriminant analysis (OPLS-DA) plot of lipid features correlated with presentation of diabetes in White (pink circle) and AA (burgundy circle) cohorts. **B)** Bar graph plot displaying scores on LV1 showing lipids that distinguish diabetes in the White cohort. Top 10 correlated features to diabetes in the White cohort are highlighted in pink. **C)** OPLS-DA plot of inflammatory features correlated with presentation of diabetes in White (pink circle) and AA (burgundy circle) groups. **D)** Bar graph plot displaying scores on LV1 which show inflammatory characteristic features of diabetes in White (pink bars) and AA (burgundy bars) groups. Top 10 correlated features to diabetes in White and AA groups are highlighted in pink and burgundy, respectively. X-axis represents scores on latent variable (LV) 1. Y-axis represents scores on LV2 not used for analysis.

Next, we analyzed inflammatory profiles using the same methodology. We observed a striking separation between Dx-AA and Dx-White with a model classification error of cross-validation = 0.3, significantly above random error (**Fig. 5C**). These results were different from what we saw when selecting inflammatory features within each racial group (**Suppl. Fig S7A and S7C).**

Consistent with what was determined in our previous univariate and multivariate analysis, cytokines MCP-1, eotaxin, and IL-27 were important for the classification of diabetes in White individuals. Importantly, we saw that TNF-α, IL-6, and IL-1β (Th1 cytokines known to induce CRP) were also important for classifying diabetes in White individuals. In contrast, IL-17A, IL-1E, IL-17F, G-CSF, and IFN-γ (Th17-associated cytokines) were important contributors to the classification of diabetes in African Americans (**Fig. 5D**). We also identified IL-33, IL-31, TGF-α, GRO-α and epithelial growth factor (EGF) (**Fig. 5D**) as important for distinguishing diabetes in African American vs White groups.

Interestingly, we also found that features characteristic of diabetes in White or African American groups were also positively correlated to disease when compared with non-disease controls within each racial population (**Suppl. Fig S7B and S7D).** Finally, TNF-α, one of the most reported markers of inflammation in diabetes, was the third most relevant feature that correlated negatively with diabetes specifically in AA (**Suppl. Fig S7D**). This reaffirms the differences seen in classical inflammation markers manifesting in African Americans, compared to White individuals. Taken together, results from this supervised clustering analysis identified systemic inflammatory features that correlated positively with the presentation of diabetes in African Americans, specifically Th17-type inflammation. Similar to the immune phenotyping results, we conclude from the OPLS-DA analysis that plasma cytokines generally reported to characterize diabetes (TNF-α, IL-6 and IL-1β) mainly do so in cases of diabetes in the White subcohort from the HANDLS study. However, this finding does not hold in the African American HANDLS subcohort.

### Relationships between lipids and inflammatory markers exhibit inverse correlations with clinical measures of diabetes in White and African American participants within the HANDLS subcohort

Our clinical findings support an interaction between lipids and inflammation as modulators of diabetes, especially in White individuals (**Fig. 2**). Literature supports the role of lipids in activation of immune cells, specially T cells and macrophages (De Jong, 2015; Hubler & Kennedy, 2016; Seufert et al., 2022). Since such immune cells could secrete much of the cytokines that were differentially modulated between racial groups upon stimulation with lipids, we next investigated the relationship between lipids and inflammatory cytokines with respect to diabetes status. To address these associations, we first calculated all possible permutations of ratios between all 128 lipids and 47 cytokines measured on a per-subject basis. This approach generated lipid:cytokine ratios. Then, we correlated each ratio with clinical markers of diabetes such as HbA1C and the homeostatic model assessment for insulin resistance (HOMA-IR), both significantly different between NoDx and Dx in both racial groups (**Fig. 2B and Suppl. Fig S8**) (Christensen et al., 2010; Khalili et al., 2023). We plotted the differences on a per-racial group basis (**Fig. 6**). We observed a striking inverse pattern in the relationships that correlated to diabetes in the White group compared to those in the African American cohort. The vast majority of lipid:cytokine ratios that significantly correlated to HbA1C (**Fig 6A**) and HOMA-IR (**Fig. 6B**) in White individuals were not significantly correlated in the African American group, and vice versa. Only a handful of ratios correlated significantly to clinical markers of diabetes in both groups, however such correlations represented weaker signals relative to the strongest race-associated ratios identified. In the White cohort, most of the ratios that correlated positively to HbA1C (**Fig 6A**) included at least one cytokine (MCP-1 or eotaxin) that was significantly increased in the White cohort in our previous analysis (**Fig. 4 and 5).** Conversely, in the African American group, relationships that included either MCP-1 and eotaxin correlated negatively to HOMA-IR (**Fig 6B**). These findings build upon previous conclusions on the cytokines that were found to be differentially increased in Dx-White over NoDx-White and Dx-AA. In African Americans, we found that the inflammatory markers soluble CD40 ligand (sCD40L) and RANTES were expressed in most of the relationships positively correlated to HOMA-IR, marker of insulin resistance (**Fig. 6B).** With this analysis, we conclude that lipid:cytokine relationships are inversely correlated to clinical markers of diabetes and insulin resistance in the White vs African American HANDLS cohort.

**Figure 6.**
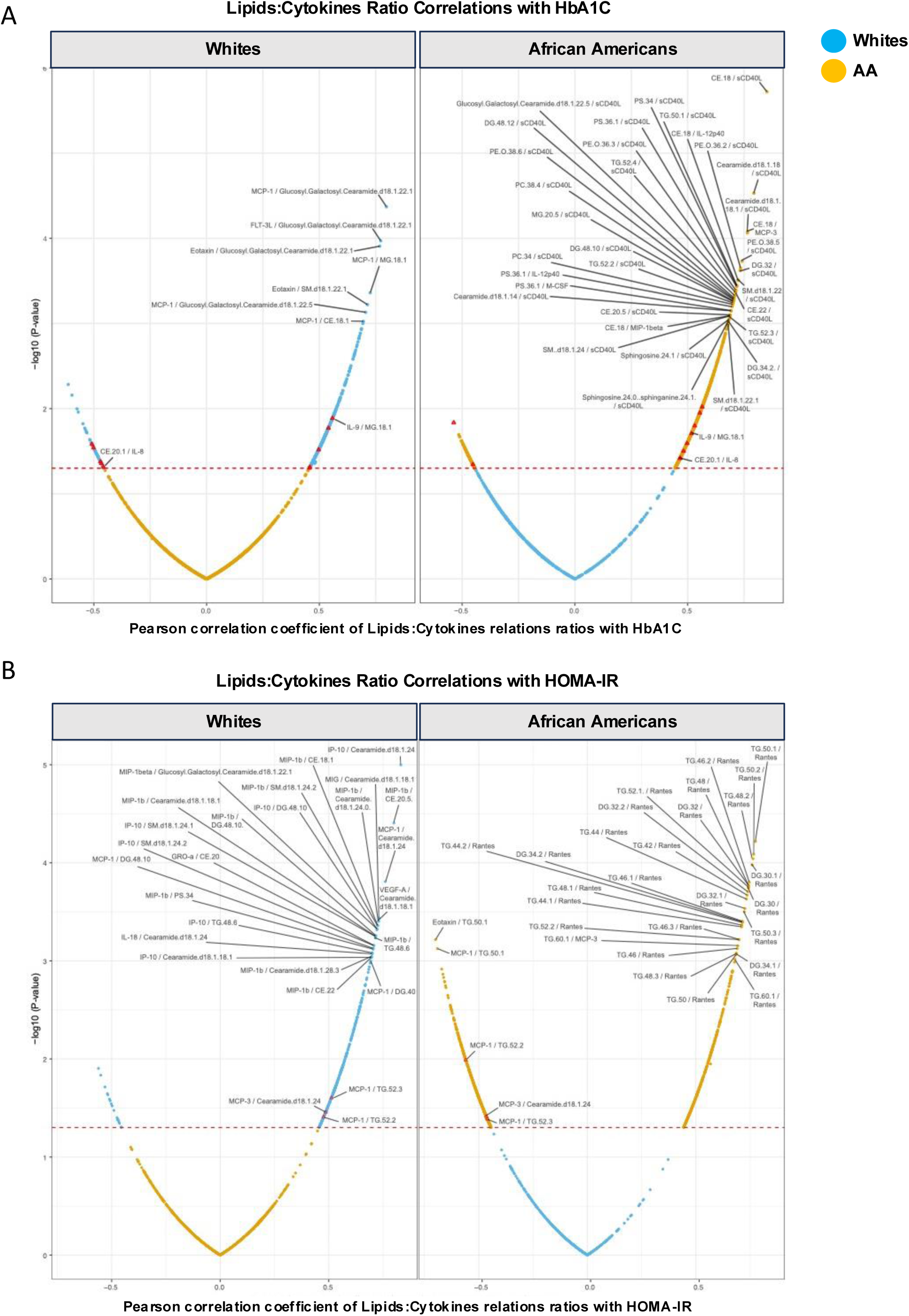
Modulatory relationships of lipids and inflammatory markers in White and AA groups correlate inversely to diabetes markers. Volcano plots showing all permutated lipid:cytokine relationships in White and AA. Briefly, all ratios were calculated for each lipid:cytokine relation, correlated to HbA1C (A) and HOMA-IR (B), and subset for all correlations that were significant in each racial group only or in both. Correlations uniquely significant in white are colored in blue and correlations uniquely significant in AA are colored in yellow . Significant correlations in both groups are represented by red triangles. X axis indicates the pearson correlation coefficient. Y axis indicates the –log10 of p-values for the lipid/cytokine relationships correlated to HbA1C (A) and HOMA-IR (B). The dotted red line represents threshold of significance values, above p-value<0.05 and below p-value>0.05.

### Lipid and inflammatory features of diabetes seen in the HANDLS subcohort are validated in a T2D subcohort from the AllofUs multi-ethnic study

Given our findings describing dramatic differences in lipids, inflammatory markers, and lipid:cytokine ratios between the White and African American groups from the HANDLS subcohort (**Fig. 6**), we wanted to determine whether we could observe similar outcomes in a larger cohort. To this end, we investigated the differences in clinical parameters associated with diabetes, dyslipidemia, and inflammation using a T2D cohort from the multi-ethnic study AllofUs (N=17,339).

By evaluating the same clinical measurements that were significantly modulated in the HANDLS diabetes subcohort in the AllofUs T2D subcohort, we noted similarities and differences (**Fig. 7**). First, we evaluated diabetes standard-of-care clinical markers, such as HbA1C, glucose, and insulin in the AllofUs T2D subcohort (**Suppl. Fig. S9**). Initially, we observed that HbA1C (**Suppl. Fig. S9A**) and insulin **(Suppl. Fig. S9C)** were significantly increased in African Americans with T2D, compared to the White group with T2D. On the other hand, glucose was not significantly different between White and African American groups with T2D **(Suppl. Fig. S9E).** When we performed a linear regression model adjusting for BMI and age, two biological variables used to match comparison groups in the HANDLS diabetes subcohort, we found that only HbA1C remained higher in African Americans with T2D (**Suppl. Fig. S9B, S9D, and S9F**). Second, similar to the HANDLS diabetes subcohort, we noticed that CholHDLRat (**Fig. 7A**) and total triglycerides (**Fig. 7C**) were significantly increased in the White population with T2D compared to African Americans with T2D. Third, opposite to the hs-CRP findings in the HANDLS diabetes subcohort, we noticed that classical inflammation detected by standard CRP levels, was significantly increased in AA with T2D, compared to White individuals with T2D (**Fig. 7E**). These findings became even more evident when we performed a linear regression model adjusting for BMI and age in the AllofUs T2D subcohort (**Fig. 7B, 7D, and 7F**).

**Figure 7.**
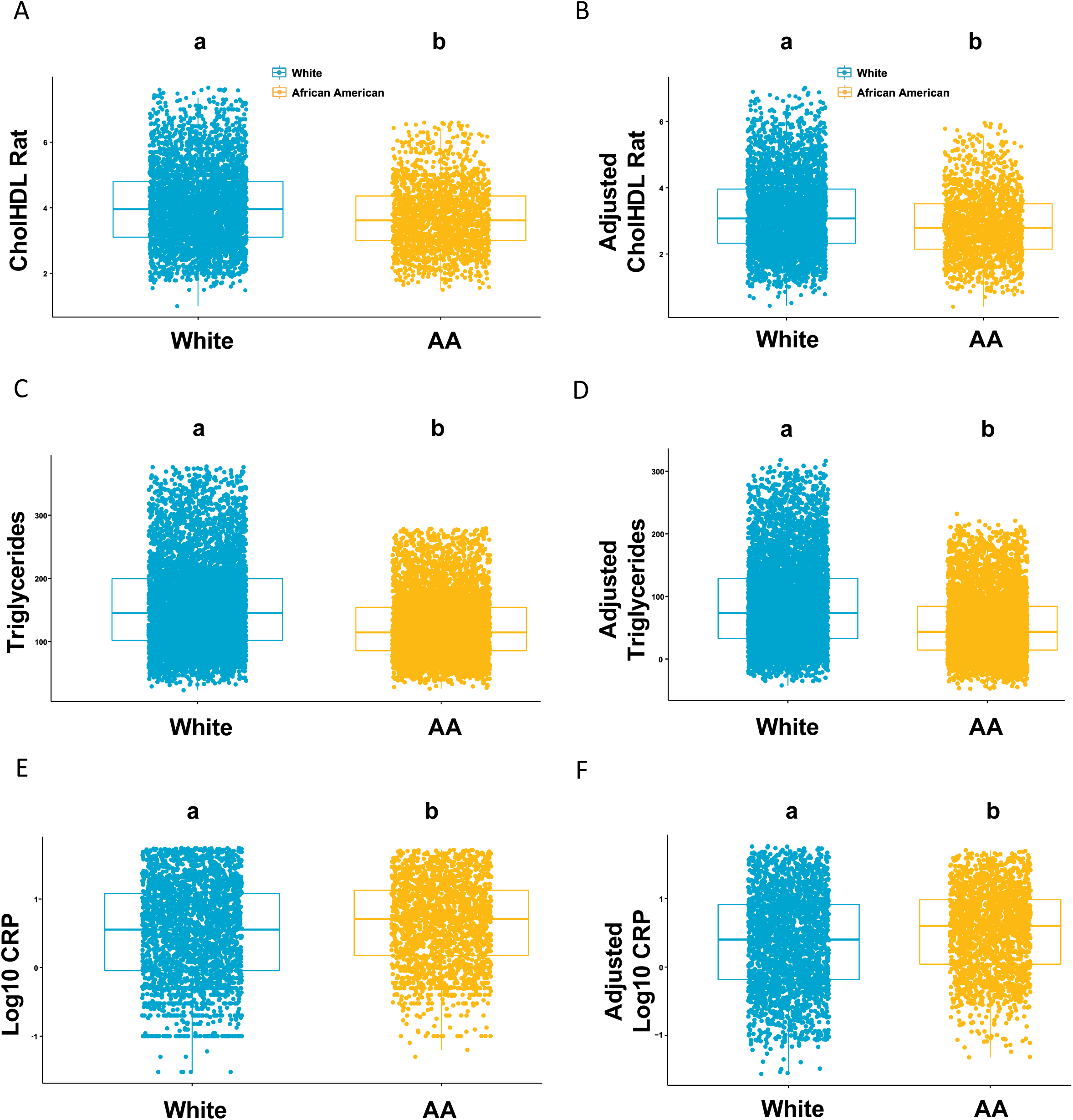
Clinical lipid and inflammatory parameters in AllofUs T2D subcohort confirm differential features seen in HANDLS diabetes subcohort. Clinical parameters that were differentially associated in White and AA groups from HANDLS subcohort were assessed using the multi-study AllofUs. Differences between the means of White and AA are plotted for CholHDLRat (A), triglycerides (C), and CRP (represented by logarithmic values due to exponential distribution) (E). A linear regression model was performed in the AllofUs T2D subcohort adjusting for variables body mass index (BMI) and age, biological variables used to match comparison groups in HANDLS diabetes subcohort. Adjusted differences between the means of Whites and AA are shown for CholHDLRat (B), triglycerides (D), and CRP (represented by logarithmic values due to exponential distribution) (F). The X axis represents race and the Y axis represents the clinical parameters evaluated. The White population is color represented in teal and the AA population is color represented in mustard. Statistical test used for comparison was Student T-test.

Overall, by using the AllofUs T2D subcohort to validate the clinical findings obtained in the analysis of our well-matched HANDLS diabetes subcohort, we confirmed that dyslipidemia distinctively characterizes diabetes in White vs African American cohorts.

## DISCUSSION

Our study demonstrates a disparity in the relationship of lipids and inflammatory mediators to indicators of glycemic control, potentially providing an explanation for how diabetes persists as a health disparity. In this study, we leveraged data from two well-described cohorts, HANDLS and AllOfUs. With HANDLS (N=40), we demonstrated that lipid profiles abundant in triglycerides, diacylglicerides, and ceramides characterize diabetes distinctively in the White group (with diagnosis of diabetes vs. without diagnosis of diabetes) compared to the African American group with diabetes. Such profiles failed to characterize diabetes in African Americans (with diagnosis of diabetes vs. without diagnosis of diabetes). Our findings also show that diabetes in the White groups is characterized by dyslipidemia associated with a classic systemic inflammatory signature. This signature can be identified using CholHDLRat, TGs, and hs-CRP as clinical proxies. Conversely, diabetes in the AA cohort is characterized by a Th17-type inflammation. Further, we show that systemic relationships between lipids and inflammatory markers in the HANDLS subcohort inversely correlate to indicators of glycemic control in each population evaluated. We validated elevated dyslipidemia in diabetes in White vs African American HANDLS participants in a large cohort of people with T2D (N=17,339) through the multi-ethnic study AllofUs.

In our study, we first conclude that clinical lipid measurements, though accounting for variability in our dataset, only reflect diabetes status in White and not in African American groups. Dysregulation of clinically measured lipids or dyslipidemia has been reported in inter-ethnic comparative studies. For example, one report suggested that minority groups, except for African Americans, are generally more likely to have high TGs and low HDL levels compared to White groups (Frank et al., 2014). Further, most research addressing dyslipidemia and disease presentation comparing diverse populations concludes that some routinely measured lipids do not reflect risk in cases of cardiovascular disease and reproductive disorders in African Americans (Koval et al., 2010; Mcintosh et al., 2013), consistent with this study.

Results from our lipidomics analysis revealed that a variety of TGs were significantly increased in diabetes in our White cohort compared to the African American cohort. These findings, specifically in the White HANDLS cohort, recapitulate previous conclusions from cohorts of European ancestry. For example, in the Malmo cohort researchers found that increased levels of TG and DG were associated with increased risk of T2D similar to our White cohort having increased TG and DG when diagnosed with diabetes (Fernandez et al., 2020). In concordance with our findings, studies in AA continuously report healthier lipid profiles in this population (Bentley & Rotimi, 2017; Frank et al., 2014; Mcintosh et al., 2013). These findings support that plasma and clinical lipids are not uniformly related to diabetes risk and disease presentation, and affirm that AA risk for type 2 diabetes may be underestimated, thereby contributing to the health disparity in disease burden in African Americans. Further, our data may explain why a racial disparity in efficacy of lipid lowering drugs to improve HbA1c persists (Cromer et al., 2023), despite lipid lowering drugs being equally if not more effective for cardiovascular risk in AA populations compared to White populations (Kalra, 2021).

Our findings suggest that the classical markers of inflammation CRP, IL-6, and TNF-α mainly discriminate diabetes from non-diabetes cases in the White HANDLS cohort, but not in the African American group. Our findings in the White cohort are consistent with increased levels of CRP, IL-6, and TNF-α observed in T2D in several studies (Bowker et al., 2020; Effoe et al., 2015; Kristiansen & Mandrup-Poulsen, 2005; Mirza et al., 2012; Swaroop et al., 2012). In our study we found that cytokines eotaxin, MCP-1, and IL-27 were increased in diabetes, though only in the White cohort and not in African American group. Eotaxin and MCP-1 are inflammatory markers reported to be elevated in patients with T2D (Okdahl et al., 2022; Panee, 2012), while IL-27 is reported as an inhibitor of Th17 cell proliferation (Hunter & Kastelein, 2012; Jouhault et al., 2023). In this study, supervised feature selection revealed that cytokines secreted by Th17 cells (IL-17F, IL-17E, and IL-21) were amongst the top features that correlated positively with diabetes in AA. This Th17-type signature in AA was accompanied by higher levels of IL-33. A role for IL-33 in modulating the balance between Th1/Th17 cells in autoimmune disorders has been postulated (Liu et al., 2019). Since Th17 type cytokines were associated with diabetes in the African American group, our findings may implicate a non-classical inflammatory mechanism contributing to diabetes presentation. Several studies support such a mechanism including the reported relationship of T2D-associated inflammation with Th17 cell cytokines (Ip et al., 2016; Nicholas et al., 2019) (Ip et al., 2016; Jagannathan-Bogdan et al., 2011) and the discovery that reduced IL-17 is associated with improved glucose management (Sumarac-Dumanovic et al., 2013). Our study reveals novel features characterizing diabetes-related inflammation distinctively in the HANDLS African American cohort. Our conclusions suggest that specific immune features reported in literature as relevant for diabetes could be impacted by the lack of diversity of the cohorts studied, hence contributing to the lack of efficacy in discovering and targeting immune pathways central to diabetes pathophysiology based on presentation.

In this study, we observed race-dependent correlations of lipid-inflammatory marker ratios to HbA1c and HOMA-IR. Specifically, we uncovered broad correlations of lipids to RANTES and CD40L ratios in the AA cohort. Though its role and mechanisms remain under debate, the chemokine RANTES, also called CCL5, is associated with T2D, glucose intolerance, and obesity (Chou et al., 2016; Dworacka et al., 2014; Yao et al., 2014). In a loss-of-function murine study, it was found that genetic deficiency of CD40L attenuated the development of diet-induced obesity, hepatic steatosis, and increased systemic insulin sensitivity (Poggi et al., 2011). Our findings correlating CD40L and RANTES to several types of lipids like PC, PE, cholesterol ester (CE), sphingomyelins (SM), and ceramides in Dx-AA could suggest the existence of an unexplored interplay among endogenous lipids, inflammation, diabetes and insulin sensitivity.

The AllofUs dataset enabled us to corroborate the lipid differences observed in the HANDLS subcohort. However, the hs-CRP findings from the HANDLS diabetes subcohort were not replicated using CRP values in the AllofUS T2D subcohort. This difference could be due to a limitation in comparison between both cohorts, given that the HANDLS study assesses hs-CRP, while the large cohort in the AllofUS data was limited to CRP levels in T2D, which is measured with a less sensitive assay. Even if this limitation does not account for the discrepancy between the HANDLS and AllofUS cohorts due to reported significant positive correlation between hs-CRP and CRP levels (Helal et al., 2012), there are several other factors that could account for the difference. Unlike the HANDLS cohort, in AllofUs we could not account for poverty status, a social determinant of health that may contribute to the elevated CRP observed in the AllofUS cohort. Literature directly implicates lower socioeconomic status in increased systemic inflammation and in increased risk of diabetes in AA (Arnold et al., 2020; Boylan et al., 2020; Butler, 2017; Cooper et al., 2024; Gaskin et al., 2014; Muscatell et al., 2020; Van Dyke et al., 2017). In this context, our data reaffirms the importance of including sociological measurements in studies evaluating health and disease in diverse populations (Williams et al., 2016).

Further limitations of our study include a relatively small sample size (N = 40, HANDLS cohort) and limited inclusion of sociological factors other than to match comparison groups. We overcame these limitations by using the AllofUs dataset to validate key clinical findings from the reduced but matched participants from the HANDLS cohort. However, the AllofUs participants could not be matched with all the same variables as in HANDLS.

In summary, we show that presentation of diabetes is heterogenous. By using comparative analysis and diverse cohorts, these results raise fundamental questions regarding how diabetes and specifically T2D is managed in the clinic based on their TG levels and CRP status. Future research addressing the efficacy of Th17 anti-inflammatory therapy, especially in patients who do not achieve glycemic control targets is warranted. Finally, our study highlights the need for large scale diabetes trials to be diverse to capture the full spectrum of disease presentation and intervention outcomes, thus paving the way for mechanistic understanding and individualized approaches to diabetes management.

## Supporting information

Manuscript Supplemental Table

## Author Contributions

Conceptualization, Gabriela Pacheco-Sanchez and Dequina Nicholas; Data curation, Gabriela Pacheco-Sanchez, Miranda Lopez, Alan B. Zonderman, Michele K. Evans, Marcus Seldin and Dequina A. Nicholas; Formal analysis, Gabriela Pacheco Sanchez, Miranda Lopez, Leandro M. Velez, Ian Tamburini, Naveena Ujagar, Julio Ayala, Gabriela De Robles, Hannah Choi, John Arriola, Rubina Kapadia, Cholsoon Jang, Marcus Seldin, and Dequina A. Nicholas; Funding acquisition, Gabriela Pacheco Sanchez, Cholsoon Jang, Marcus Seldin, and Dequina A. Nicholas; Methodology, Gabriela Pacheco Sanchez, Miranda Lopez, Leandro M. Velez, Ian Tamburini, Naveena Ujagar, Julio Ayala, Gabriela De Robles, Alan B. Zonderman, Michele K. Evans, Cholsoon Jang, Marcus Seldin, Dequina A. Nicholas; Project administration, Rubina Kapadia, Alan B. Zonderman, Michele K. Evans, Cholsoon Jang, Marcus Seldin, and Dequina A. Nicholas; Supervision, Gabriela Pacheco Sanchez, Cholsoon Jang, Marcus Seldin, and Dequina A. Nicholas; Writing – original draft, Gabriela Pacheco Sanchez, Miranda Lopez, Hannah Choi, John Arriola, Dequina A. Nicholas; Writing – review & editing, Gabriela Pacheco Sanchez, Miranda Lopez, Leandro M. Velez, Ian Tamburini, Naveena Ujagar, Julio Ayala, Gabriela De Robles, Hannah Choi, John Arriola, Rubina Kapadia, Alan B. Zonderman, Michele K. Evans, Cholsoon Jang, Marcus Seldin, and Dequina A. Nicholas.

## Data Availability Statement

Not applicable.

## Supplemental Figures

**Supplemental Figure S1.**
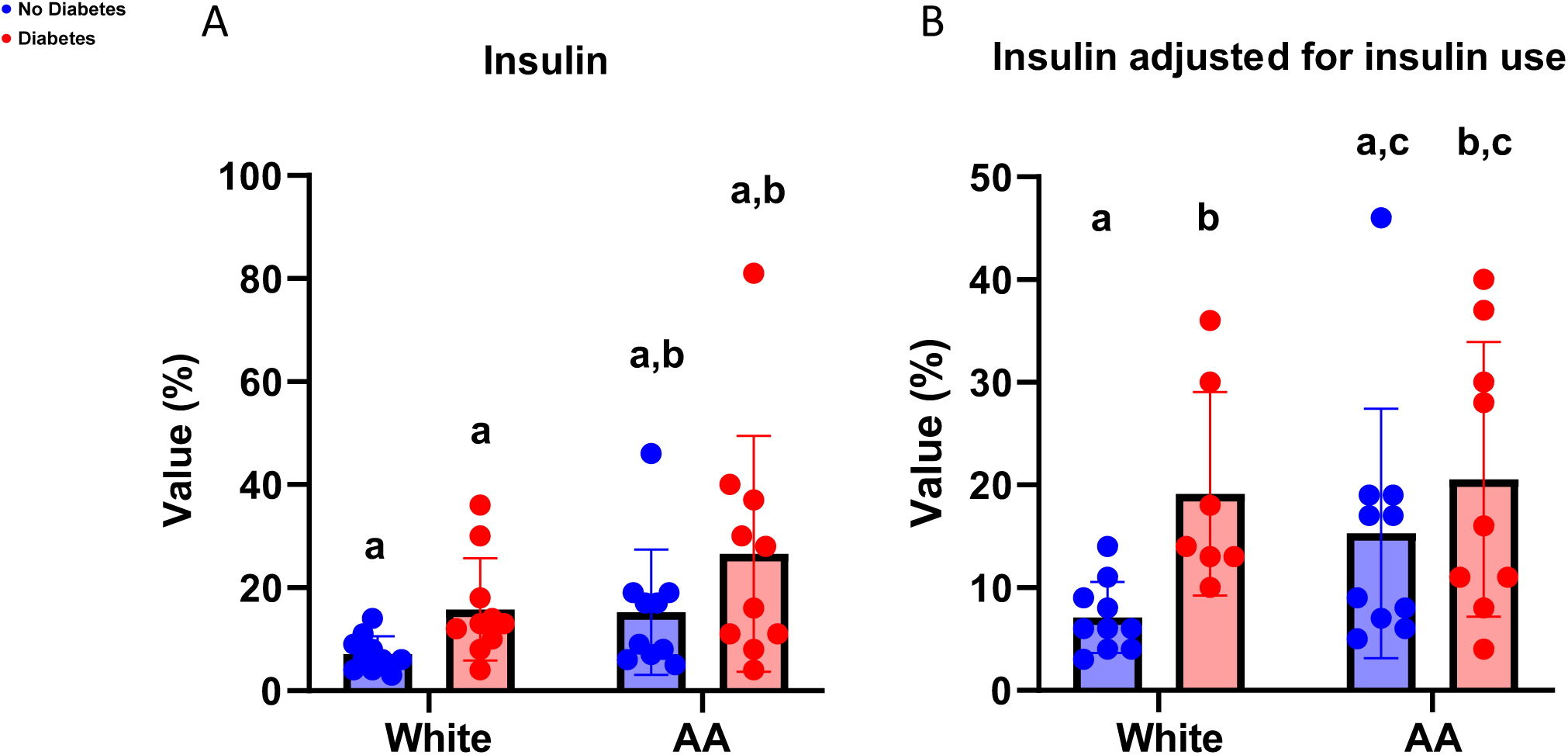
Statistical post-ANOVA comparison of insulin. Bar/dot graph showing results from multiple statistical comparisons of insulin levels among Whites and AA without and with diabetes. Insulin levels for all participants**(A)** and adjusted for participants who were not prescribed insulin **(B)** are shown . Blue = People without diabetes, red = people with diabetes. Statistical analysis performed using Two-way ANOVA with followed by Fisher’s LSD post-comparison test (unadjusted p-values). Statistical post-ANOVA comparisons were performed only between matching groups based on diabetes status and race and comparisons between groups with and without diabetes were excluded from the analysis. P-values obtained from analysis were represented using a statistical letter system, where significantly different p-values are represented by different letters and non-significant p-values are represented by same letters.

**Supplemental Figure S2.**
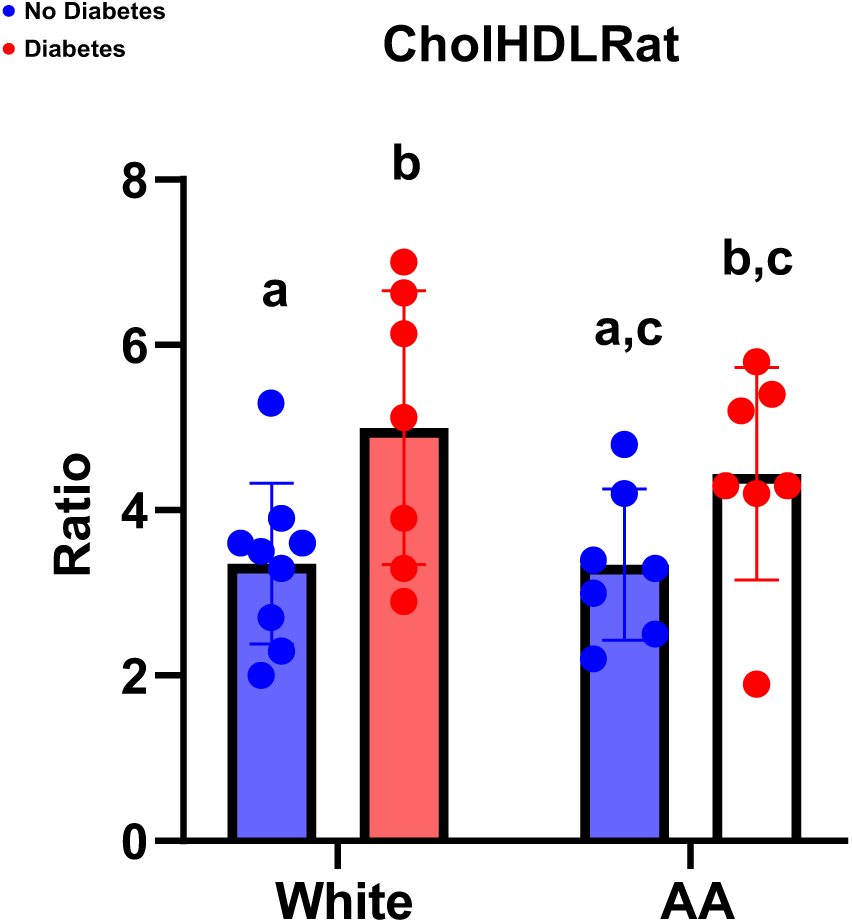
Statistical post-ANOVA comparison of Cholesterol/HDL ratio (CholHDLRat) adjusting for statins (lipid lowering drug) use. Bar/dot graph showing results from multiple statistical comparison of CholHDLRat among Whites and AA without and with diabetes. Blue = People without diabetes, red = people with diabetes. Statistical analysis performed using Two-way ANOVA with followed by Fisher’s LSD post-comparison test (unadjusted p-values). Statistical post-ANOVA comparisons were performed only between matching groups based on diabetes status and race and comparisons between groups with and without diabetes were excluded from the analysis. P-values obtained from analysis were represented using a statistical letter system, where significantly different p-values are represented by different letters and non-significant p-values are represented by same letters.

**Supplemental Figure S3.**
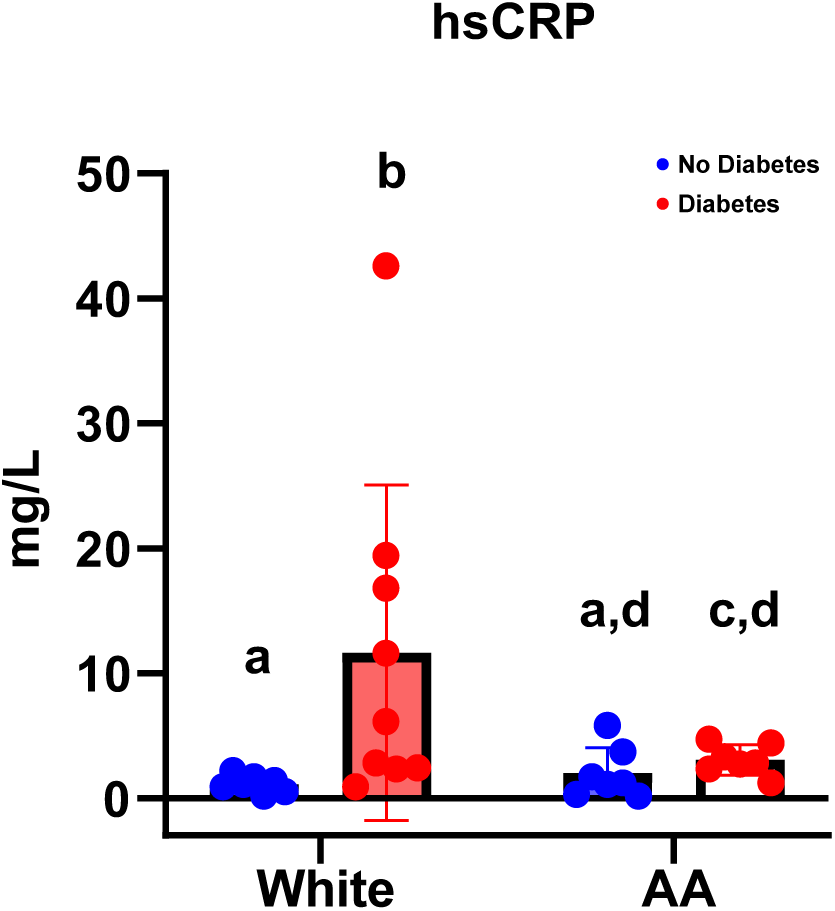
Statistical post-ANOVA comparison of high sensitivity C-reactive protein (hsCRP) adjusting for statins (lipid lowering drug) use. Bar/dot graph showing results from multiple statistical comparison of hsCRP among Whites and AA without and with diabetes. Blue = People without diabetes, red = people with diabetes. Statistical analysis was performed using Two-way ANOVA with followed by Fisher’s LSD post-comparison test (unadjusted p-values). Statistical post-ANOVA comparisons were performed only between matching groups based on diabetes status and race and comparisons between groups with and without diabetes were excluded from the analysis. P-values obtained from analysis were represented using a statistical letter system, where significantly different p-values are represented by different letters and non-significant p-values are represented by same letters.

**Supplemental Figure S4.**
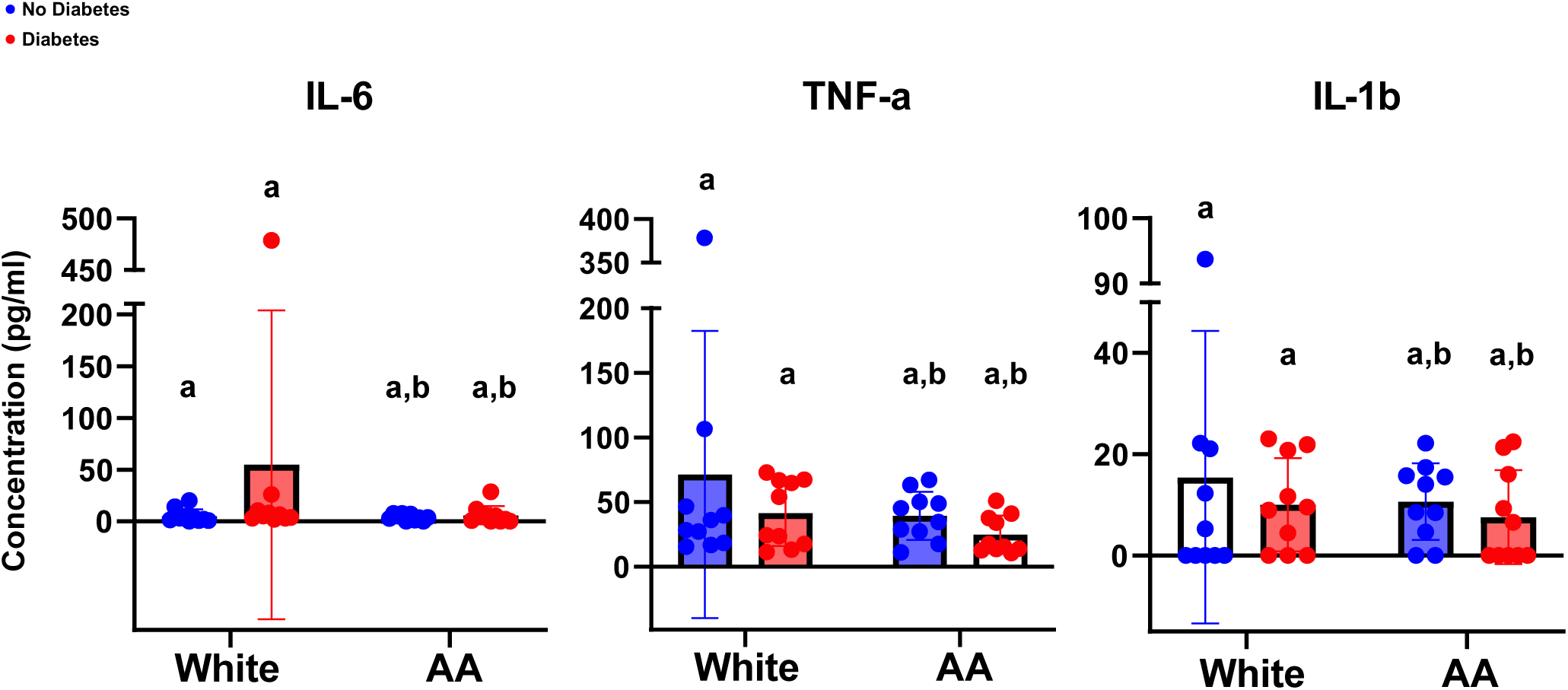
Statistical post-ANOVA comparison of classical inflammatory markers IL-6, TNF-a, and IL-1b in HANDLS subcohort. Bar/dot graph showing results from post-ANOVA multiple statistical comparisons among Whites and AA without and with diabetes for IL-6 (left), TNF-a (middle), and IL-1b (right). Blue = People without diabetes, red = people with diabetes. Statistical analysis was performed using Two-way ANOVA with followed by Fisher’s LSD post-comparison test (unadjusted p-values). Statistical post-ANOVA comparisons were performed only between matching groups based on diabetes status and race and comparisons between groups with and without diabetes were excluded from the analysis. P-values obtained from analysis were represented using a statistical letter system, where significantly different p-values are represented by different letters, and non-significant p-values are represented by the same letters.

**Supplemental Figure S5.**
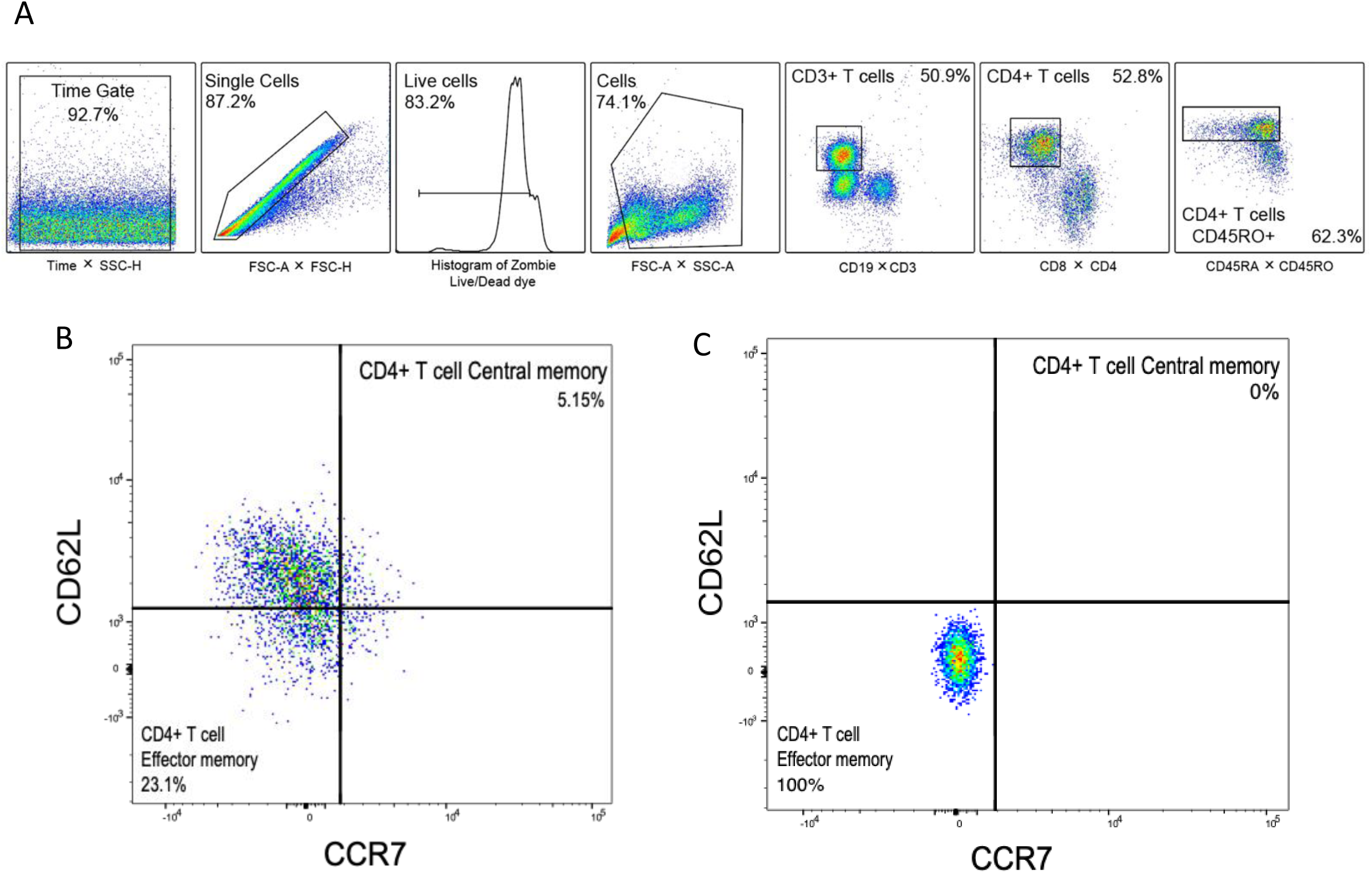
Gating strategy for CD4+ Central Memory T cells. **A)** First, cells were gated by time of sample acquisition followed by doublet discrimination using FCS-H and FSC-A. Next live cells were gated based on viability dye. Then for size and granularity for lymphocytes and myeloid cells based on SSC-A and FSC-A. After that CD3+ T cells were gated based on CD3 and CD19 antibodies. From CD3+ T cells, CD4+ T cells were gated based on antibodies CD4 and CD8. Next, CD4+CD45RO+ T cells were gated based on CD45RO and CD45RA antibodies. **B)** Lastly, CD4+ Central memory T cells were gated based on markers CCR7 and CD62L. **C)** Stain controls were included in the experiment to verify positive populations. Percentages on plot represent frequencies based on parent population.

**Supplemental Figure S6.**
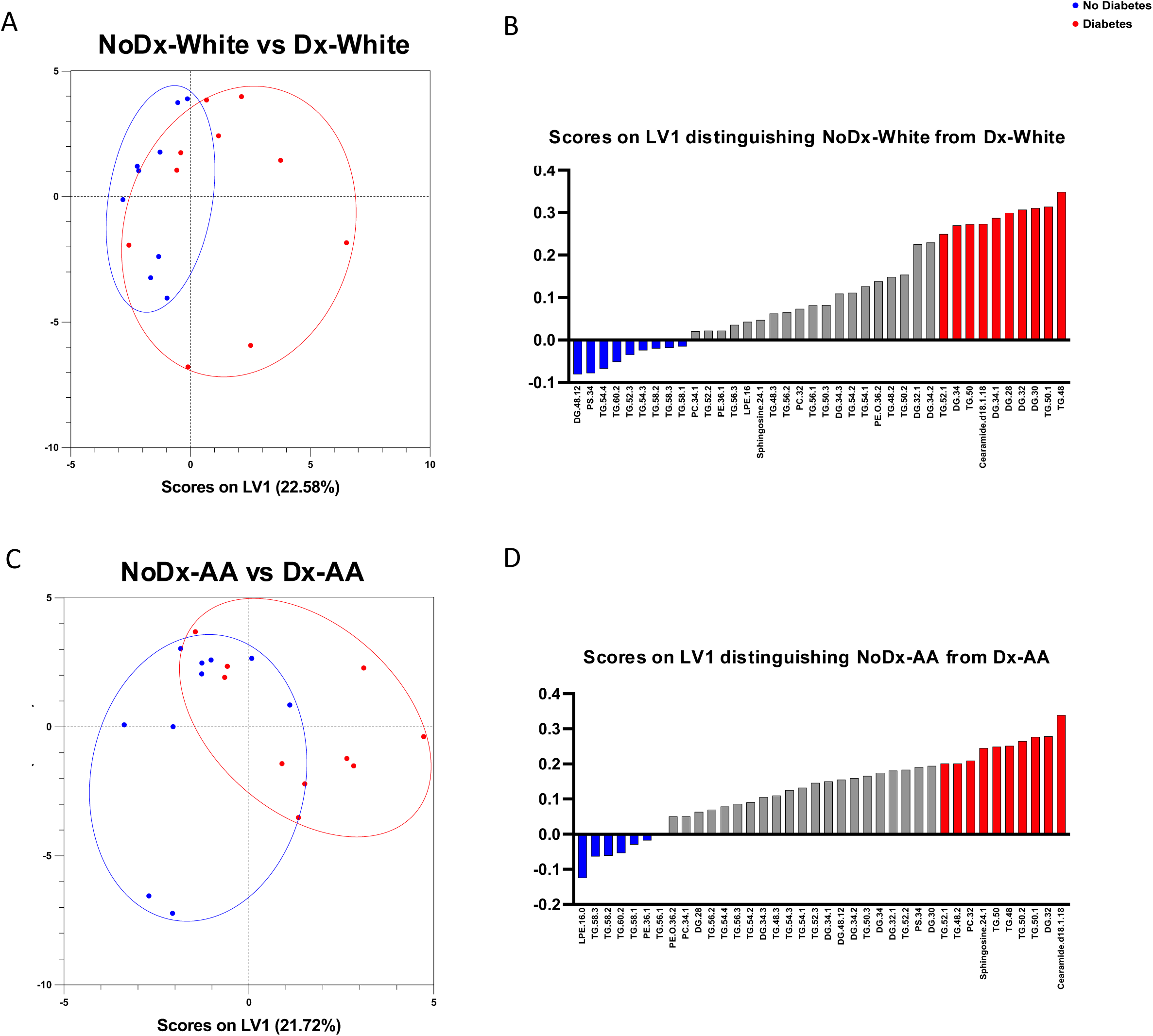
Lipid features characteristics of White and AA with diabetes in a diverse HANDLS subcohort. Orthogonal partial least squares discriminant analysis (OPLS-DA) plot of lipids features correlated with presentation of diabetes in White **(A)** and AA **(C)**. Blue dots refer to group without diabetes and red dots refer to group with diabetes. X-axis represents scores on latent variable (LV) 1 and Y-axis represent scores on LV2 not used for analysis. Bar graph plot displaying scores on LV1 showing lipids that distinguish diabetes in White **(B)** and AA **(D)**. Top 10 features that correlated positively (red) and negatively (blue) to diabetes in White and AA are colored in the charts.

**Supplemental Figure S7.**
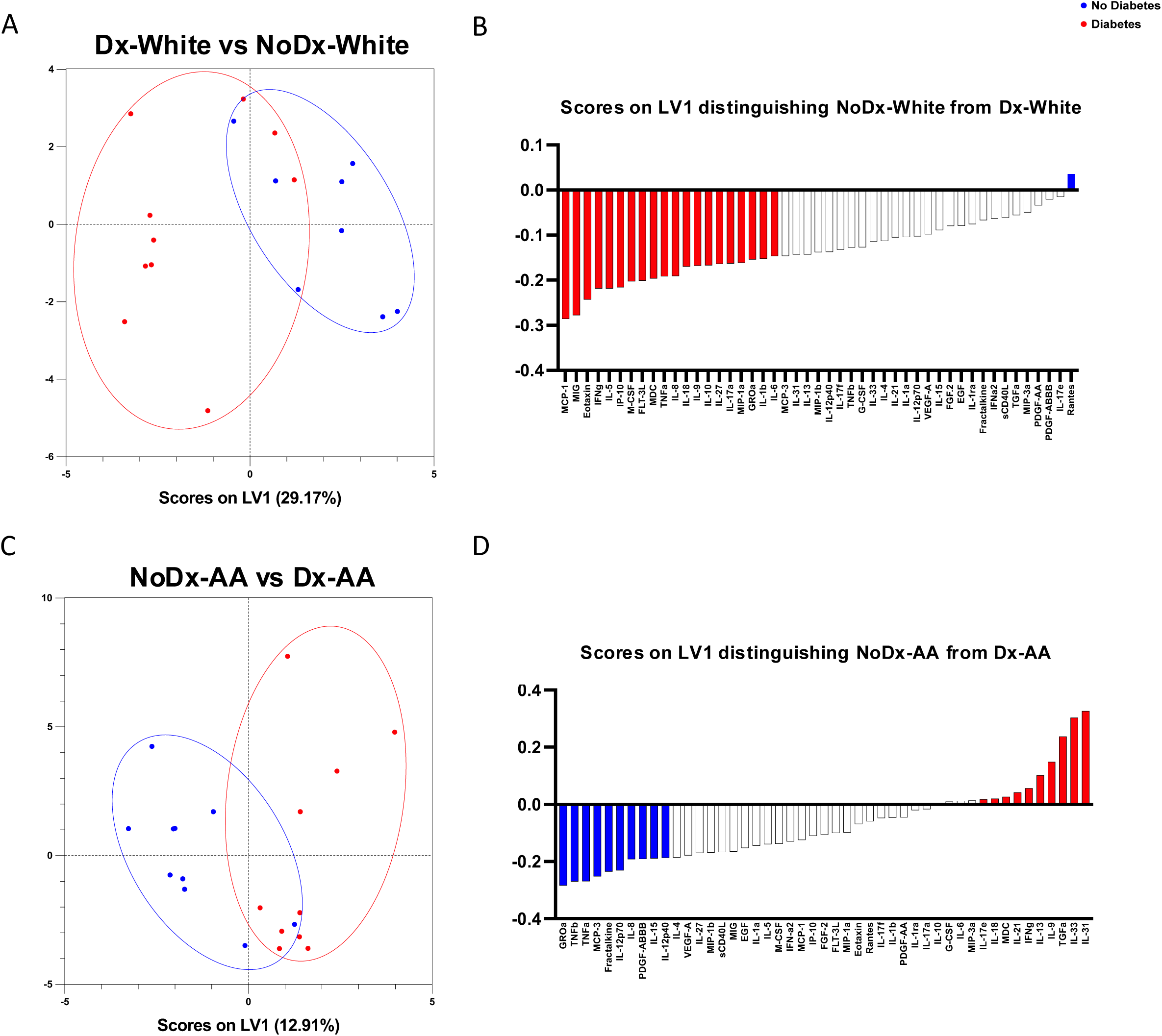
Inflammatory features characteristics of White and AA with diabetes in a diverse HANDLS subcohort. Orthogonal partial least squares discriminant analysis (OPLS-DA) plot of lipids features correlated with presentation of diabetes in White **(A)** and AA **(C)**. Blue dots refer to participants without diabetes and red dots refer to participants with diabetes. X-axis represents scores on latent variable (LV) 1 and Y-axis represents scores on LV2 not used for analysis. Bar graph plot displaying scores on LV1 showing lipids that distinguish diabetes in white **(B)** and AA **(D)**. Top 10 features that correlated positively (red) and negatively (blue) to diabetes in white and AA are colored in the charts.

**Supplemental Figure S8.**
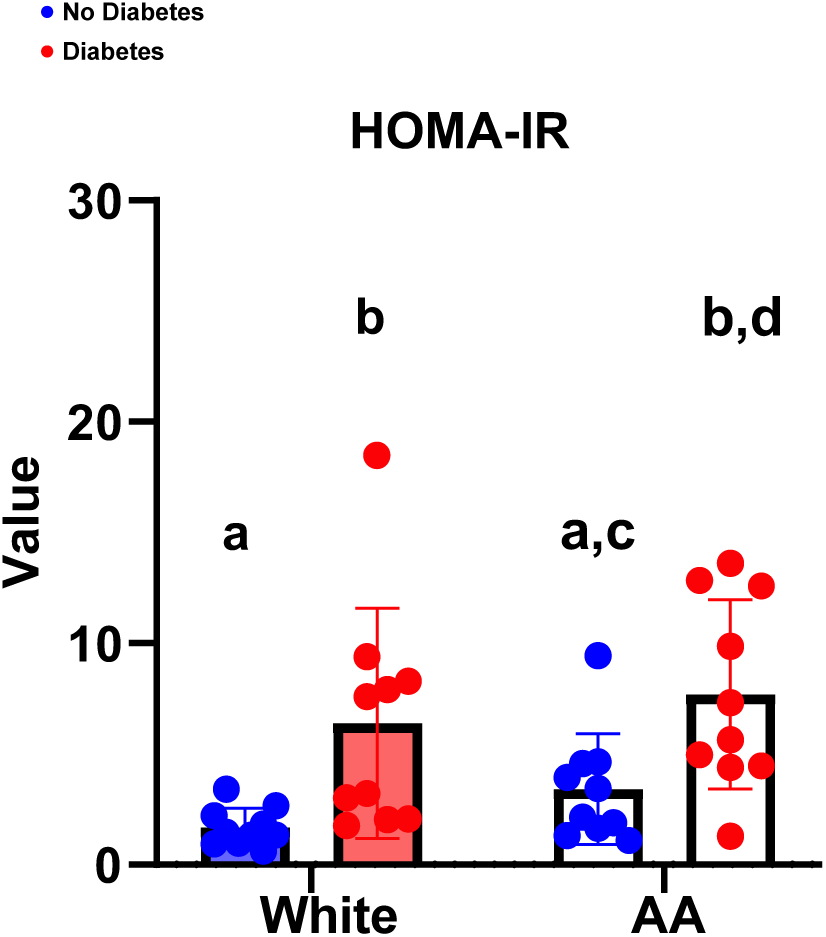
Statistical post-ANOVA comparison of homeostasis model assessment for insulin resistance (HOMA-IR). Bar/dot graph showing results from multiple statistical comparisons of HOMA-IR values among Whites and AA without and with diabetes. Blue = People without diabetes, red = people with diabetes. Statistical analysis performed using Two-way A NOVA with followed by Fisher’s LSD post-comparison test (unadjusted p-values). Statistical post-ANOVA comparisons were performed only between matching groups based on diabetes status and race and comparisons between groups with and without diabetes were excluded from the analysis. P-values obtained from analysis were represented using a statistical letter system, where significantly different p-values are represented by different letters and non-significant p-values are represented by same letters.

**Supplemental Figure S9.**
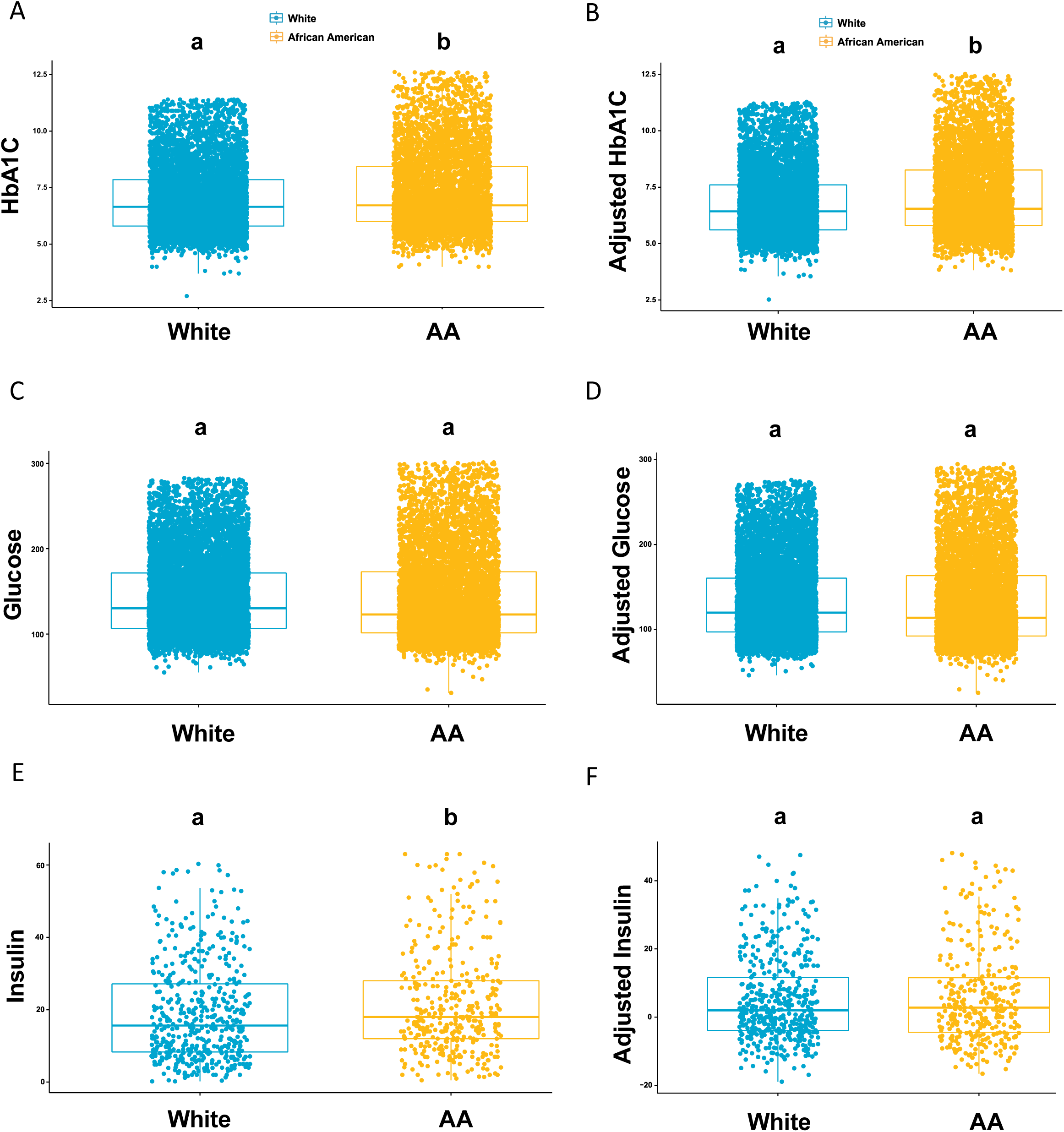
Clinical lipid and inflammatory parameters in AllofUs T2D subcohort confirm differential features seen in HANDLS diabetes subcohort. Clinical parameters that were differentially associated to diabetes in socially diverse populations from HANDLS subcohort were assessed using the multi-study AllofUs. Differences between the means of white and AA are plotted for HbA1C **(A)**, glucose **(C)**, and insulin **(E)**. A linear regression model was performed in the AllofUs T2D subcohort adjusting for variables body mass index (BMI) and age, biological variables used to match comparison groups in HANDLS diabetes subcohort. Adjusted differences between the means of whites and AA are shown for HbA1C **(B)**, glucose **(D)**, and insulin **(F)**. X axis represent race of group and Y axis represents clinical parameters evaluated. White population are colored represented in teal and AA population are colored represented in mustard. Statistical test used for comparison was student T-test.

