## Supplementary material for "Comparative analysis of White and African American groups reveals unique lipid and inflammatory features of diabetes": Manuscript Supplemental Table

*These authors contributed equally

### SUPPLEMENTARY TABLES:

**Supplementary Table S1.** List of dietary intake lipids and statistical differences (p-values) Using Two-way ANOVA and post-ANOVA tests comparing all four groups in the HANDLS subcohort (N=40)

**Table legend:** A subcohort of 40 individuals from the HANDLS study were divided into 4 comparison groups based on disease status and race: White without diabetes (NoDx-White), White with diabetes (Dx-White), African Americans without diabetes (NoDx-AA), and African Americans with diabetes (Dx-AA). Values were transformed using Box-Cox transformation and comparisons were made using the two-way Anova test with post-anova comparisons using the Fisher LSD’s test. P-values are displayed as decimal numbers.

| Dietary Intake type of Food | ANOVA Overall | Race | Diabetes | Interaction of race x diabetes | NoDx-White vs Dx-White | NoDx-AA vs Dx-AA | Dx-White vs Dx-AA | NoDx-White vs NoDx-AA |
|---|---|---|---|---|---|---|---|---|
| lfa4 | 0.4839 | 0.3267 | 0.2271 | 0.9484 | 0.4160 | 0.3663 | 0.5154 | 0.4586 |
| lfa6 | 0.3427 | 0.2655 | 0.1794 | 0.5927 | 0.5611 | 0.1855 | 0.6783 | 0.2452 |
| lfa8 | 0.5159 | 0.3589 | 0.2359 | 0.9343 | 0.3683 | 0.4326 | 0.4787 | 0.5532 |
| lfa10 | 0.6516 | 0.5139 | 0.2897 | 0.8088 | 0.3574 | 0.5605 | 0.5272 | 0.7706 |
| lfa12 | 0.6113 | 0.3092 | 0.5564 | 0.5206 | 0.3854 | 0.9692 | 0.2425 | 0.7882 |
| **lfa14 *** | **0.0256** | **0.0072** | 0.2884 | 0.2887 | 0.9996 | 0.1364 | 0.2175 | **0.0086** |
| lfa16 | 0.4393 | 0.6300 | 0.1855 | 0.4045 | 0.7224 | 0.1296 | 0.8018 | 0.3534 |
| lfa16_1 | 0.4580 | 0.9928 | 0.5767 | 0.1350 | 0.4991 | 0.1477 | 0.2896 | 0.2840 |

| | | | | | | | | |
|---|---|---|---|---|---|---|---|---|
| Ifa18 | 0.3228 | 0.4286 | 0.1309 | 0.4526 | 0.5816 | 0.1118 | 0.9769 | 0.2773 |
| Ifa18_1 | 0.6823 | 0.8135 | 0.3078 | 0.5396 | 0.7707 | 0.2499 | 0.5483 | 0.7888 |
| Ifa18_2 | 0.7162 | 0.7022 | 0.5078 | 0.3874 | 0.8850 | 0.2823 | 0.3787 | 0.7313 |
| Ifa18_3 | 0.7610 | 0.7125 | 0.4071 | 0.5705 | 0.8517 | 0.3249 | 0.8877 | 0.5087 |
| Ifa18_4 | 0.6983 | 0.8370 | 0.2598 | 0.7708 | 0.3159 | 0.5510 | 0.7253 | 0.9516 |
| Ifa20_1 | 0.4208 | 0.4525 | 0.2745 | 0.3055 | 0.9607 | 0.1373 | 0.2115 | 0.8442 |
| Ifa20_4 | 0.0946 | 0.7932 | 0.2647 | 0.0244 | 0.3951 | **0.0187** | 0.0728 | 0.1489 |
| Ifa20_5n3 | 0.6478 | 0.3145 | 0.6068 | 0.55444 | 0.4352 | 0.9565 | 0.2604 | 0.7664 |
| Ifa22_1 | 0.2885 | 0.6881 | 0.4275 | 0.0868 | 0.5024 | 0.0783 | 0.3441 | 0.1345 |
| Ifa22_5n3 | 0.1117 | **0.0188** | 0.5527 | 0.8809 | 0.5990 | 0.7530 | 0.0731 | 0.1112 |
| Ifa22_6n3 | 0.5814 | 0.2157 | 0.5645 | 0.8135 | 0.5660 | 0.8093 | 0.2965 | 0.4743 |
| MonoFat | 0.6792 | 0.8269 | 0.3225 | 0.4974 | 0.8236 | 0.2403 | 0.5259 | 0.7441 |
| PolyFat | 0.6928 | 0.7249 | 0.4988 | 0.3569 | 0.8606 | 0.2606 | 0.3685 | 0.6849 |
| Saturated Fat | 0.3251 | 0.2889 | 0.2182 | 0.3604 | 0.8186 | 0.1320 | 0.9161 | 0.1653 |
| Fat | 0.5599 | 0.4762 | 0.6840 | 0.1554 | 0.9196 | 0.1605 | 0.5873 | 0.4383 |
| Carbohydrates | 0.4358 | 0.4762 | 0.6840 | 0.1554 | 0.4666 | 0.1964 | 0.6084 | 0.1335 |
| Total sugar | 0.4229 | 0.3679 | 0.5996 | 0.1926 | 0.1973 | 0.5760 | 0.7703 | 0.1220 |
| Protein | 0.1265 | 0.5151 | 0.8180 | **0.0.233** | 0.1394 | 0.0741 | 0.2340 | **0.0392** |
| Energy | 0.4654 | 0.5553 | 0.5393 | 0.1797 | 0.5998 | 0.1683 | 0.5880 | 0.1735 |
| Dash Saturated Fat | 0.5206 | 0.5399 | 0.2238 | 0.5399 | 0.1976 | 0.6643 | 0.3873 | >0.9999 |
| Dash Total Fat | 0.8178 | 0.3922 | 0.7271 | 0.8130 | 0.6790 | 0.9365 | 0.6598 | 0.4399 |
